# Investigating the link between depressive symptoms and resting-state brain connectivity in people with breast cancer: A systematic review

**DOI:** 10.1101/2025.09.19.25336219

**Authors:** Jacqueline B Saward, Annalee L Cobden, Priscilla Gates, Juan F Dominguez, Karen Caeyenberghs

**Affiliations:** Cognitive Neuroscience Unit, School of Psychology Deakin University, Geelong, Australia; School of Psychological Sciences and Turner Institute for Brain and Mental Health, Monash University, Melbourne, Australia; Department of Health Services Research, Peter MacCallum Cancer Centre. Parkville, Victoria, Australia

**Keywords:** Systematic review, breast cancer, depressive symptoms, functional connectivity, resting state fMRI

## Abstract

**Purpose:** Depressive symptoms are a common and debilitating experience among people with breast cancer (BC), often impacting quality of life and recovery. However, the neural mechanisms underlying these symptoms are unclear. This systematic review synthesised resting-state functional magnetic resonance imaging (rsfMRI) literature in BC populations to identify functional connectivity (FC) correlates of depressive symptoms.

**Methods:** A systematic search of EMBASE, PsycINFO, Medline, and CINAHL identified 27 eligible studies (15 cross-sectional, 12 longitudinal) examining the relationship between depressive symptoms and functional connectivity using rsfMRI in BC participants. Data were extracted on study design, participant characteristics, depressive symptoms, imaging acquisition, FC outcomes, and reported associations. Study quality was assessed using the Newcastle-Ottawa Scale. Findings were synthesised qualitatively.

**Results:** Across cross-sectional studies, BC participants showed elevated depressive symptoms and widespread FC alterations, predominantly patterns of dysconnectivity, compared to healthy controls. However, most studies did not find significant associations between depressive symptoms and FC. Longitudinal studies revealed dynamic trajectories in depressive symptoms and FC patterns with cancer treatment or training intervention.

**Conclusion:** While depressive symptoms are frequently reported by BC participants, the underlying neural mechanisms remain unclear, possibly due to methodological and participant heterogeneity across studies.

**Implications for cancer survivors:** Findings highlight the importance of timely and ongoing monitoring of depressive symptoms across the cancer care continuum. Future research should conduct more sensitive assessments of depressive symptomatology (e.g., ecological momentary assessment), adopt standardised rsfMRI protocols, and apply integrative network analysis. These future studies will inform rsfMRI metrics to be used as biomarkers to guide treatment at the individual cancer patient level.

## Introduction

An estimated 30% of people with breast cancer (BC) report experiencing depressive symptomatology, with prevalence rates approximately two to three times higher than the general population [1]. Depressive symptoms encompass a constellation of affective (e.g., low mood, loss of interest or pleasure), cognitive (e.g. sense of worthlessness or guilt, concentration difficulties, thoughts of death or suicide), and somatic disturbances (e.g., fatigue, psychomotor agitation or slowing, sleep issues, appetite or weight changes) [2]. Among people with breast cancer, depressive symptoms often emerge or intensify in response to the cumulative impacts of the disease and its treatments, which typically include surgery, chemotherapy, radiation, and hormonal therapy, and may persist for months or even years after treatment completion [3]. Such symptoms are frequently associated with reduced treatment compliance, worsened prognosis, and an increased risk of mortality [4]. Further, depressive symptoms can adversely impact on the quality of life, daily activities, work productivity, and social participation of people with BC [3,5]. These effects often extend beyond the individual, to the caregivers and the wider support system [6]. This has led prominent international cancer institutes to promote cancer-related depressive symptoms as a high-priority research and intervention area [3].

Ample studies have examined depressive symptomatology in people with BC [1]. For example, a previous meta-analysis [77] of 33 studies found that BC participants (1-98 months post-treatment) reported significantly higher levels of depressive symptoms compared to healthy controls (HC), with a medium-to-large effect size (Hedges’ g = 0.83). A more recent meta-analysis [1] of 72 studies across 30 countries reported an estimated pooled prevalence of depressive symptoms in BC participants of 32.2%. In addition, the authors of this paper highlighted that 16 different assessment scales were used to measure depressive symptoms, with outcomes analysed using different analysis techniques such as mean group comparisons and proportion of participants above clinical cut-offs [1]. For example, a cross-sectional study using the Hospital Anxiety and Depression Scale (HADS) found a larger proportion of BC participants with mild (BC: 10.6%; HC: 4.9%) and severe (BC: 3.7%; HC: 1.1%) depressive symptoms compared with healthy controls ten years post-diagnosis [8]. Longitudinal studies further highlight that depressive symptoms are a significant cancer survivorship issue, while symptoms may decline over time for some people with BC, a subset may experience persistently elevated or worsening symptoms after treatment [9]. In a large longitudinal study (*n =* 4803 BC participants) [10], distinct depressive symptom trajectories were identified across the three years following diagnosis: over 70% of participants exhibited stable low or improving symptoms, 20% experienced gradual worsening, and approximately 7% showed either persistently high or delayed-onset depression.

Previous reviews have suggested that depressive symptoms in people with BC may be related to neurobiological changes in the brain [11]. Resting-state functional magnetic resonance imaging (rsfMRI) is a non-invasive technique increasingly used in BC populations to uncover alterations in communication between brain regions [12]. Resting-state fMRI provides measures on the functional synchronicity or “connectivity” (FC) of large-scale networks (e.g., the default mode network, salience network) as well as key regions within these networks, while participants are at rest (i.e., not engaged in an active task or responding to external stimuli) [12,13]. The absence of a task in rsfMRI offers practical advantages by placing fewer demands on participants and reducing performance variability (compared to task-related fMRI), making it particularly suitable for clinical populations [13]. The first study to fully leverage rsfMRI in people with BC, reported alterations in global network organisation and reduced regional FC in frontal, temporal, and striatal regions in BC participants who had received chemotherapy compared to healthy controls [14].

Since that initial study, a growing number of cross-sectional studies have documented FC alterations in BC participants, with both decreased and increased connectivity reported across diverse brain regions and networks compared to healthy controls (e.g., [15–18]). At the regional level, alterations have been observed in specific brain regions, such as decreased connectivity in the dorsolateral prefrontal cortex and inferior frontal gyrus in non-chemotherapy BC participants compared with healthy controls [15]. Notably, another study [16] included both a cancer control group (non-chemotherapy) and a healthy control group and found that only BC participants who had received chemotherapy showed altered connectivity in prefrontal and parietal regions. The inclusion of a cancer control group is essential to delineate chemotherapy-specific effects by accounting for confounding cancer-related factors such as diagnosis, surgery, radiation, and hormonal therapy [17]. Alterations in FC have also been observed across large-scale networks. For example, decreased connectivity within the default mode network has been reported in chemotherapy-treated BC participants compared with healthy controls [18]. Longitudinal studies of BC participants exposed to cancer treatment have also provided insights into the temporal dynamics of FC alterations [19,20]. For example, one study [19] reported decreased FC within the default mode and frontoparietal networks in BC participants one week after chemotherapy, with partial normalisation at six months. In contrast, another study [20] observed no longitudinal changes in FC across similar networks from post-treatment to six-month follow-up, suggesting stabilisation.

Several rsfMRI studies in people with BC have also investigated whether functional connectivity alterations are associated with health outcomes, including depressive symptoms (as reviewed in this paper [14,16,19,21–45]). Specifically, one study found a strong positive correlation between HADS scores and thalamus-postcentral gyrus connectivity (*r* = .71, *p* < .001), suggesting that increased connectivity was linked to higher depressive symptom severity [37]. Another study reported a weak negative correlation between PHQ scores and left caudate connectivity (*r* = -.20 to -.30, *p* < .05), indicating that decreased connectivity was linked to higher symptom severity [22]. Despite these emerging findings, no systematic review to date has synthesised rsfMRI studies examining depressive symptoms-functional connectivity relationships in people with BC. Clarifying this literature is essential for identifying potential biomarkers of depressive symptoms that could guide development of targeted interventions.

In this systematic review, rsfMRI studies in people with breast cancer (BC) are synthesised that report findings on depressive symptoms in relation to functional connectivity. To capture the breadth of findings across the cancer experience, studies were included that involved BC participants at various phases of survivorship, including those newly diagnosed, undergoing treatment, and post-treatment. The review has two primary aims. First, to qualitatively synthesise findings from both cross-sectional studies (including those that examined group differences or assessed a single cohort at one time point) and longitudinal studies (including both those that track changes over time following cancer treatment and training interventions) in people with BC. Studies of interest investigated (i) self-reported depressive symptoms, (ii) functional connectivity outcomes, and (iii) the associations between them. The second aim of this study was to critically evaluate key methodological and conceptual challenges in the rsfMRI literature in BC populations.

## Methods

### Search strategy

The current review was conducted in accordance with the Preferred Reporting Items for Systematic Reviews and Meta-analysis (PRISMA) 2020 guidelines [46]. The completed PRISMA checklist is available in Supplementary Material (Table S1). Systematic literature searches were conducted across four electronic databases: EMBASE, PsycINFO, Medline and CINAHL. Search terms included alternative keywords and subject headings for ‘breast cancer’ and ‘resting-state fMRI’. Full search strategies for each database are provided in Supplemental Table S2. The final search was conducted on 13^th^ August 2025. Forward citation tracking via Google Scholar and backward reference checking of included studies were conducted to identify further eligible studies.

### Eligibility criteria

Studies were eligible for inclusion if they met all the following criteria: (1) original peer-reviewed research; (2) utilised rsfMRI on a cohort of adults diagnosed with breast cancer and/or receiving or having completed cancer treatment; (3) measured depressive symptoms using a validated assessment scale; (4) examined the relationship between depressive symptoms and resting-state brain connectivity; and (5) employed either a cross-sectional or longitudinal study design. Exclusion criteria were as follows: (1) articles published in a language other than English; (2) animal studies; (3) abstracts, editorial letters, case reports, or review articles; and (4) qualitative studies only.

### Study selection procedure

Records were imported into EndNote, where duplicates were removed. Next, two reviewers (JS and AC) independently screened all titles and abstracts for relevance using Rayyan, with blinding enabled. Full-text articles of potentially eligible studies were retrieved and evaluated in detail against the inclusion and exclusion criteria. A third reviewer (KC) independently reviewed the final set of included studies to confirm eligibility. Disagreements were resolved through discussion. No machine learning or AI tools were used during selection.

### Data extraction

One reviewer (JS) extracted data, which was independently checked by two reviewers (KC and JD). Any discrepancies were discussed and resolved collaboratively. The following information was extracted from each study: (a) author and year of publication; (b) participant characteristics (e.g., sample size, mean age ± SD, breast cancer stage, cancer treatment details); (c) study design and primary clinical outcome(s) of interest; (d) depressive symptom assessment (i.e., measure used, mental health-related eligibility criteria, clinical cut-off score); (e) rsfMRI acquisition parameters (e.g., scanner type, repetition time [TR], acquisition time [TA], number of volumes, voxel size, eye condition); and (f) rsfMRI analysis techniques used to derive FC metrics; (g) reported findings on depressive symptoms and FC; and (h) correlations and other reported associations between depressive symptom scores and FC in BC populations (e.g., strength, direction, implicated brain regions or networks).

All extracted data were systematically entered into structured Excel spreadsheets to facilitate cross-study comparison and synthesis. Information relevant to quality appraisal (e.g., recruitment methods, outcome measures, follow-up) was also recorded to support the subsequent risk of bias assessment. Missing or unclear data were recorded and reported accordingly.

### Risk of bias

The quality of studies was assessed independently by two reviewers (JS and KC) using the Newcastle-Ottawa Scale (NOS) [47]. The NOS is a review tool for evaluating risk of bias of observational studies. Two versions of the NOS were applied according to the study design (cross-sectional or longitudinal). The scale evaluates three domains: selection (e.g., participant recruitment and sample representativeness), comparability (e.g., control of confounders), and outcome (e.g., reliability of outcome measures, rigour of statistical analysis, and completeness of follow-up). Each domain includes specific criteria, with selected items modified to align with the objectives of this review and to reflect the participant and methodological characteristics unique to the included rsfMRI studies (see Supplementary Table S3a and S3b for modified criteria and detailed scoring information). To rate the quality of studies, stars were awarded based on the fulfillment of the criteria within each domain. These scores were then converted into quality ratings of ‘good’, ‘fair’, or ‘poor’ based on threshold criteria adapted from guidelines from the Agency for Healthcare Research and Quality (AHRQ) and are consistent with approaches used in previous systematic reviews [48].

### Qualitative Synthesis

Due to large heterogeneity in study participant characteristics and methodologies, a meta-analysis was not feasible, hence a qualitative synthesis was conducted. Studies were first grouped by design (cross-sectional or longitudinal). Longitudinal studies were further classified by focus (cancer treatment or behavioral/cognitive training interventions). Outcomes were categorised by comparison type, direction of group differences (e.g., higher vs lower depressive symptom scores, higher vs lower values of FC metrics), longitudinal changes within the BC group, and/or group-by-time interactions. Functional connectivity findings were additionally categorised according to the brain regions or networks implicated. Frequency counts were used to identify recurring patterns across studies. Studies with overlapping participants were retained if they addressed distinct aims or employed different analytic approaches; participant data were not duplicated. This review was not prospectively registered, and no formal protocol was prepared.

## Results

### Study Selection

The PRISMA flow diagram showing the study selection process is presented in Figure 1. Of the 796 records identified through database searches, 27 rsfMRI studies [14,16,19,21–45] met the eligibility criteria and were included in the qualitative synthesis. Of these, fifteen studies (56%) were cross-sectional, and 12 studies (44%) were longitudinal, with seven examining changes pre- and post-cancer treatment (mostly chemotherapy), and five assessing changes pre- and post-training interventions (e.g. mindfulness). The full selection procedure and reasons for exclusion are provided in Supplementary Table S4.

**Figure 1.**
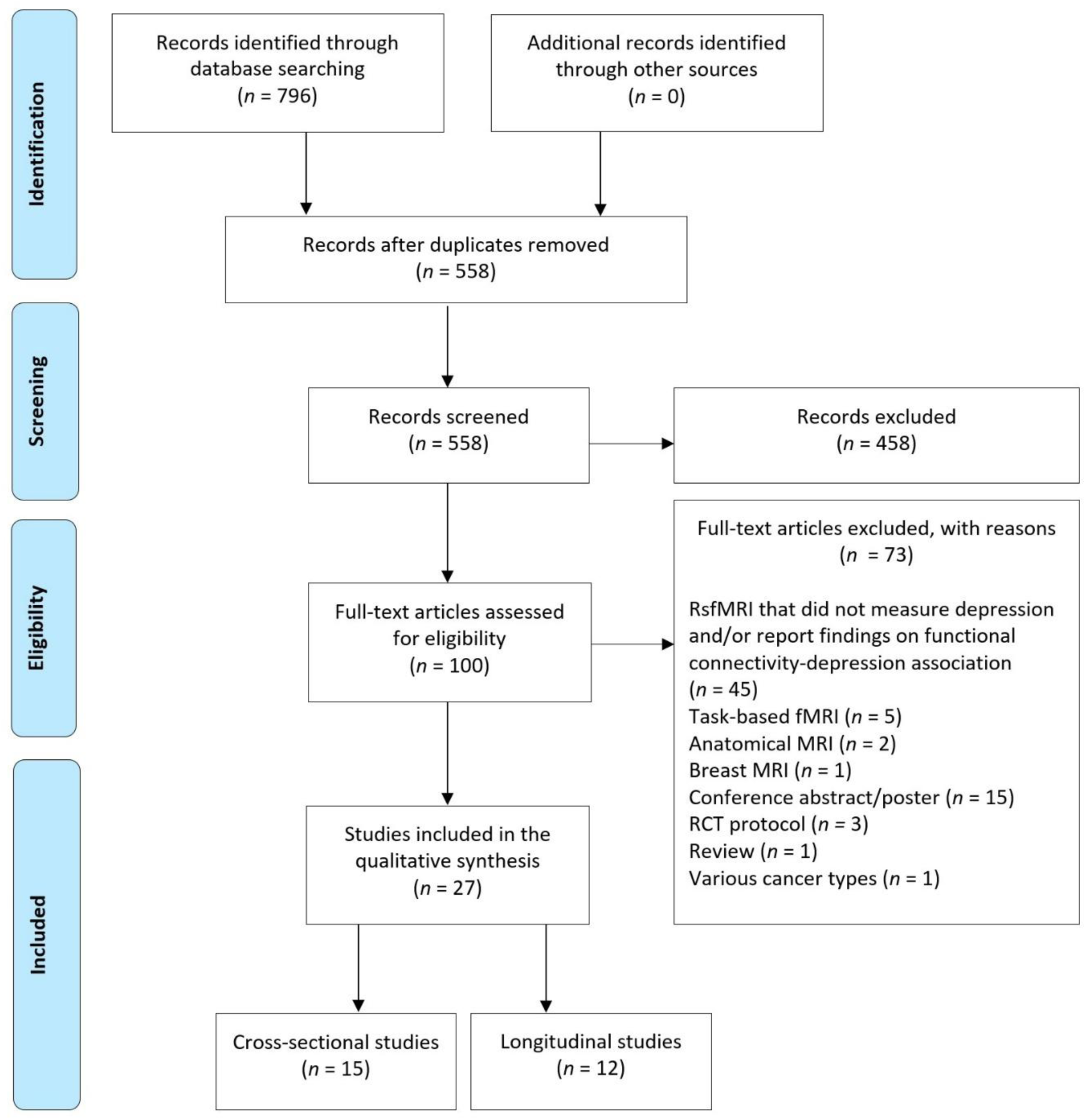
PRISMA flow diagram of the study selection process

### Risk of bias

Figure 2 presents the risk of bias for quality assessment of the studies included in this review (see Supplementary Tables S3a-S3b for modified criteria and individual study scores). None of the studies received the highest possible NOS score (9 stars), with the majority (24/27 studies, 89%) identified as having some risk of bias. After converting NOS results to AHRQ standards, 4 studies (15%) were rated as good quality (low risk of bias), 17 studies (63%) as fair quality (moderate risk of bias), and 6 studies (22%) as poor quality (high risk of bias). Further details on study quality patterns across individual NOS/AHRQ criteria can be found in Supplementary Table S4.

**Figure 2.**
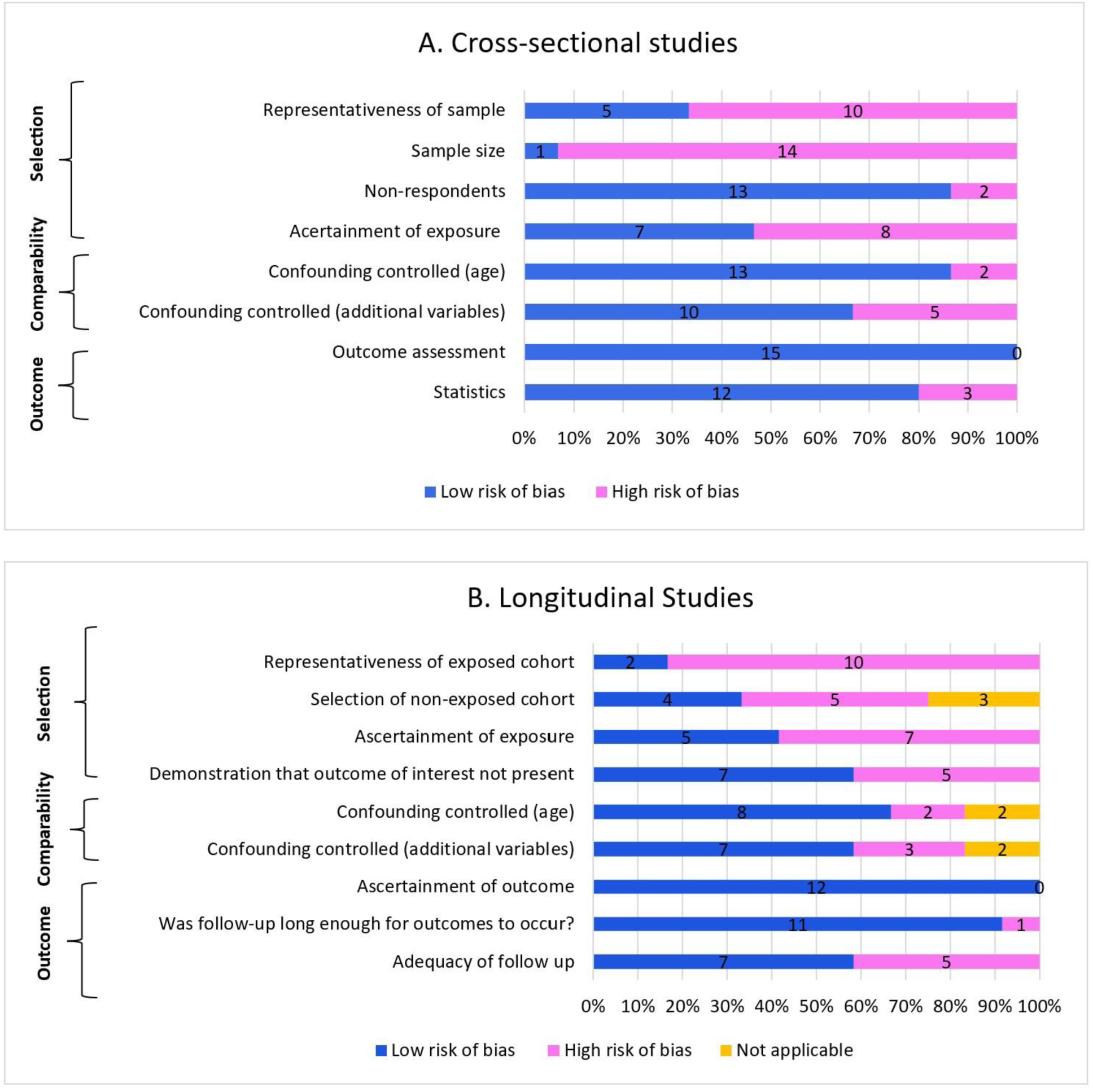
Risk of bias ratings based on the Newcastle-Ottawa Scale (NOS) *Note*. Panel A shows NOS quality ratings for cross-sectional studies; Panel B displays ratings for longitudinal studies. Numbers represent the count of studies meeting each NOS subscale criterion across selection, comparability, and outcome domains. Of note, three longitudinal studies included only a single cohort [25,36,42]. Due to the absence of a comparison or control group, these studies were rated as not applicable on the “selection of non-exposed cohort” and “control for confounding” subscale criteria but were still considered to carry a risk of bias.

### Participant characteristics

As shown in Table 1 and Supplementary Table S5, the reviewed rsfMRI studies included participants that varied widely in their clinical characteristics and in how comparison groups were structured. A total of 1,423 BC participants and 560 healthy controls were examined across the 27 studies included in the review. The majority of studies (21/27, 78%) recruited middle-aged BC participants, with mean ages over 45 years (ranging from 40.3 to 63.6). Non-metastatic breast cancer (stage 0-III) accounted for most of the BC population investigated (23/27 studies, 85%). There was large variability in cancer treatments received by BC participants, with most studies (20/27, 74%) providing incomplete details. Comparison group configurations ranged from people with BC versus healthy controls and/or cancer controls, to BC subgroups (e.g., those with/without cognitive impairment), to single BC cohorts. Participant characteristics are described in more detail in Supplementary Table S4.

**Table 1.**
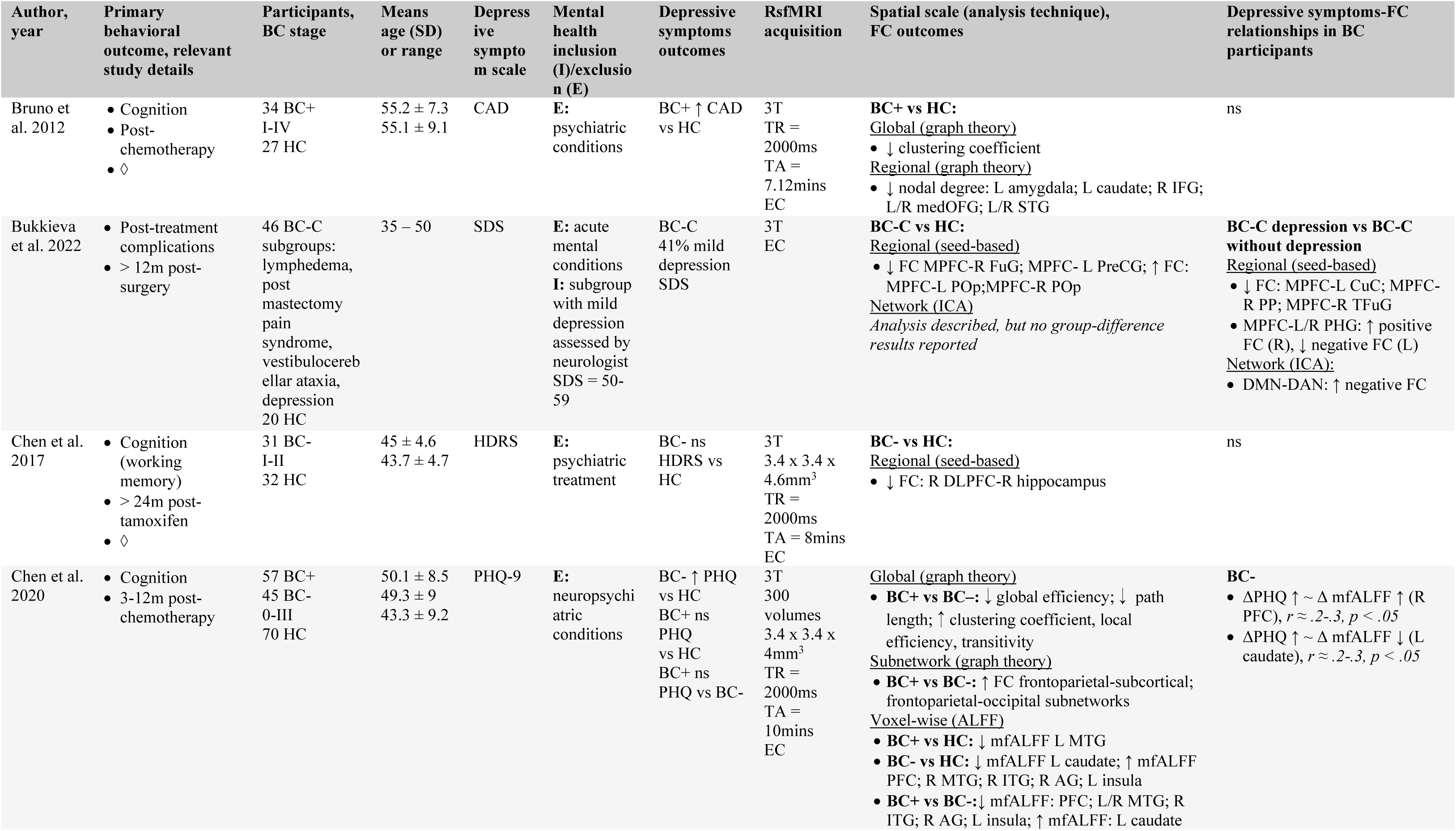
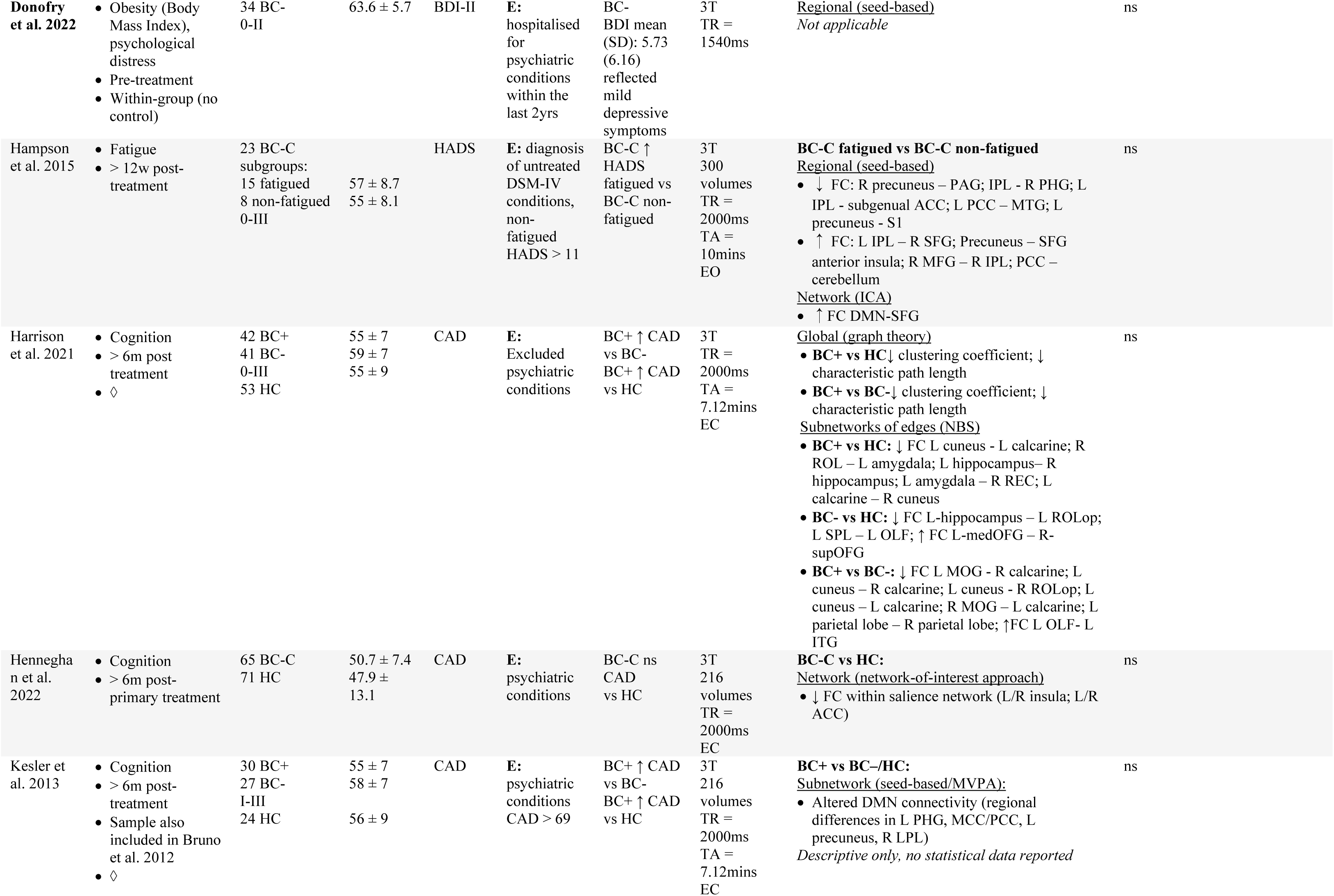
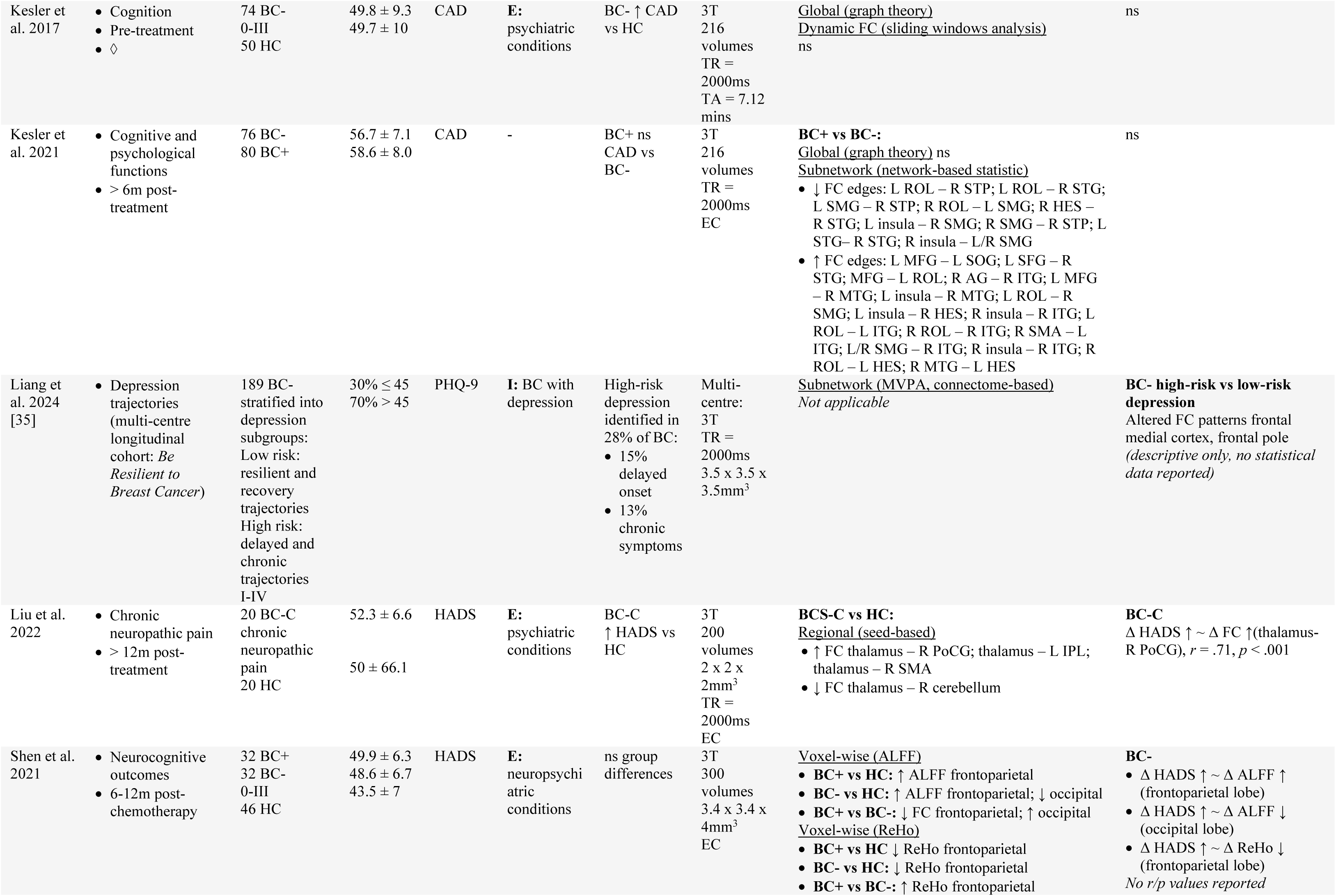
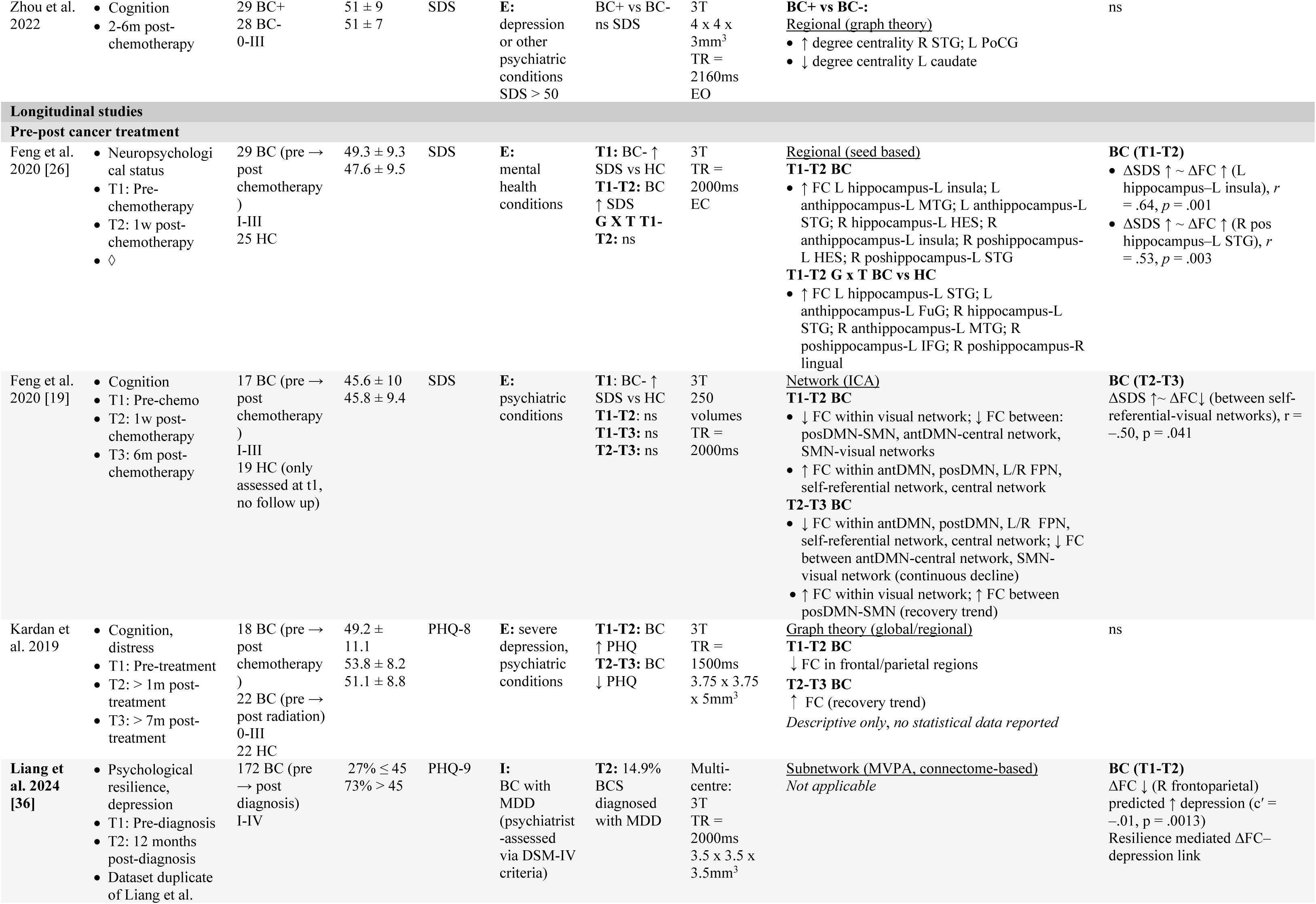
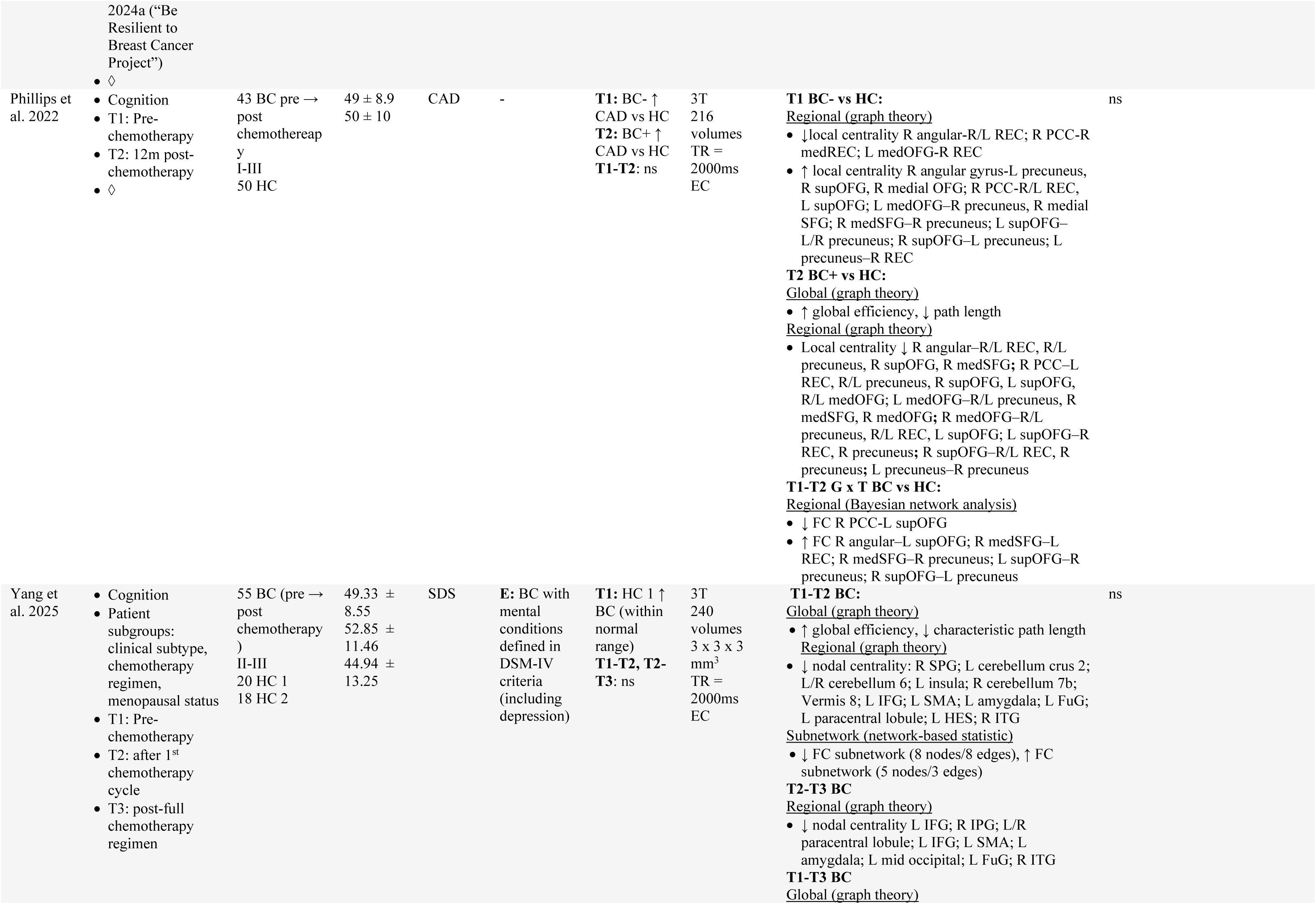
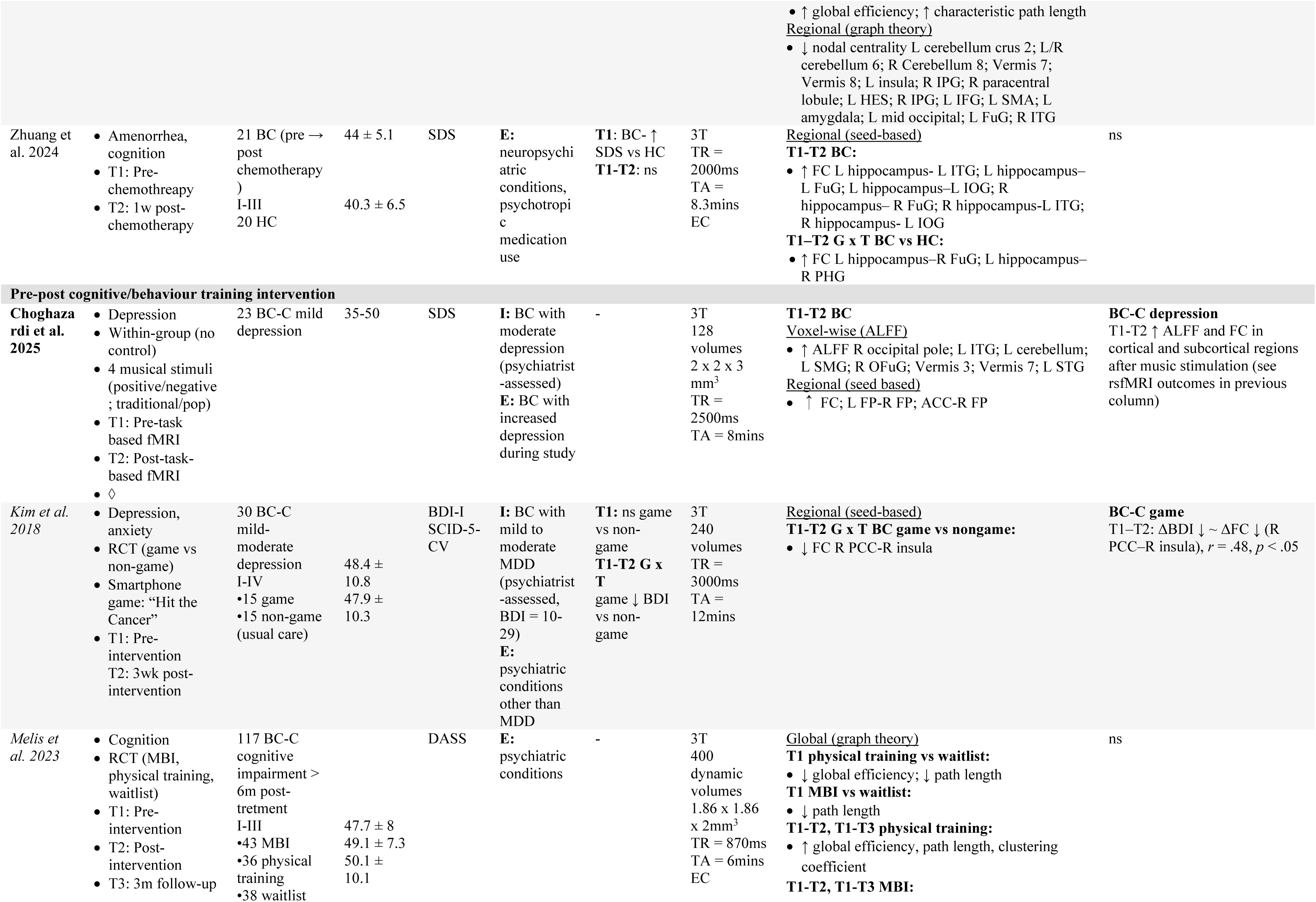
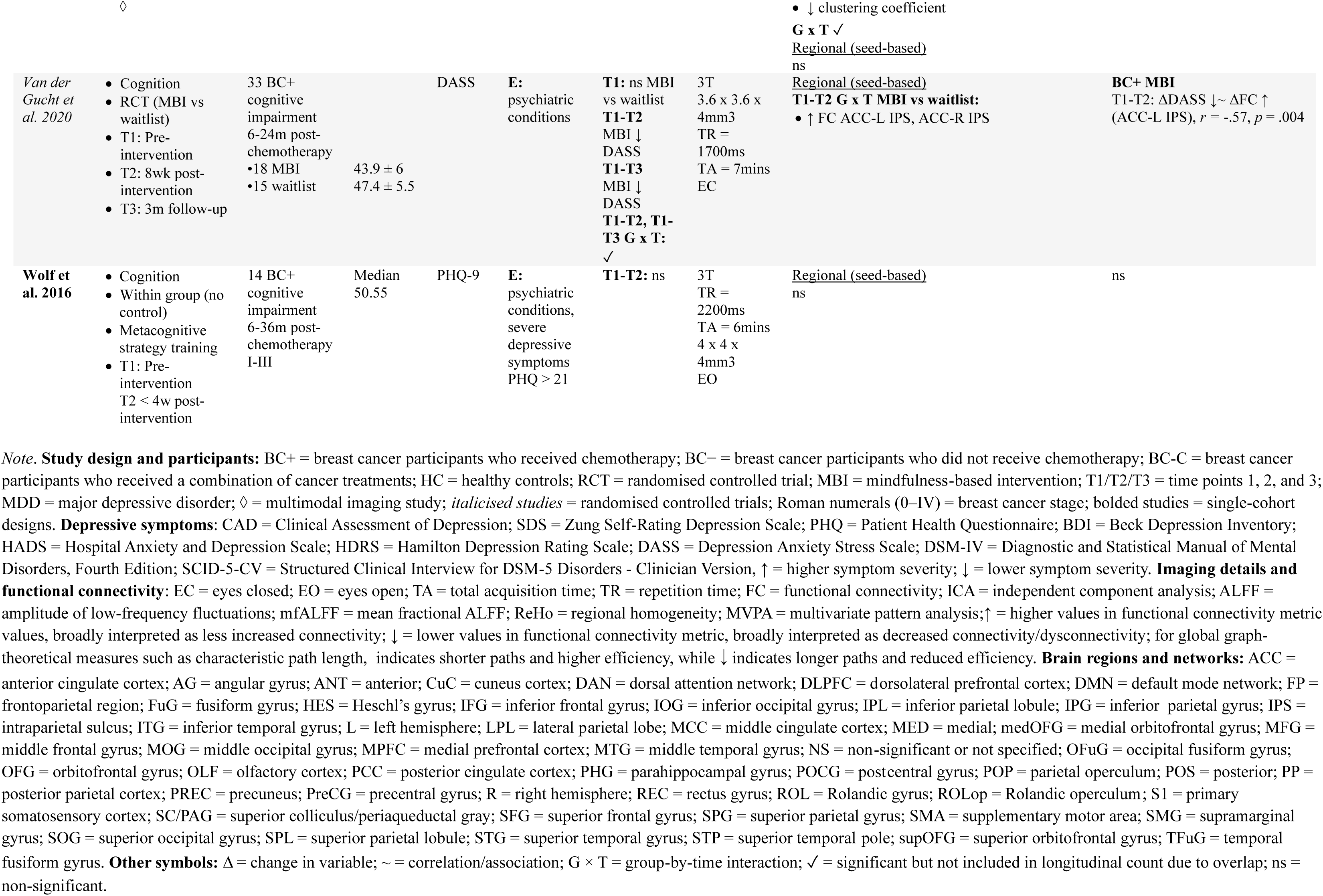
Overview of the included studies participant details, methods, and depressive symptom and functional connectivity results.

### Depressive symptoms outcomes

Table 1 and Figure 3 provide an overview of the depressive symptom measures and main findings across studies. Most studies (19/27, 70%) administered a broad battery of neuropsychological assessments, including a range of objective cognitive tests (e.g., Stroop test, digit span task) and/or self-report questionnaires (e.g., Pittsburgh Sleep Quality Index), one of which included a depressive symptom scale. Seven different depressive symptom scales were employed, with the most frequently utilised being the Clinical Assessment of Depression (CAD, 7/27 studies) and the Zung Self-Rating Depression Scale (SDS, 7/27 studies).

**Figure 3.**
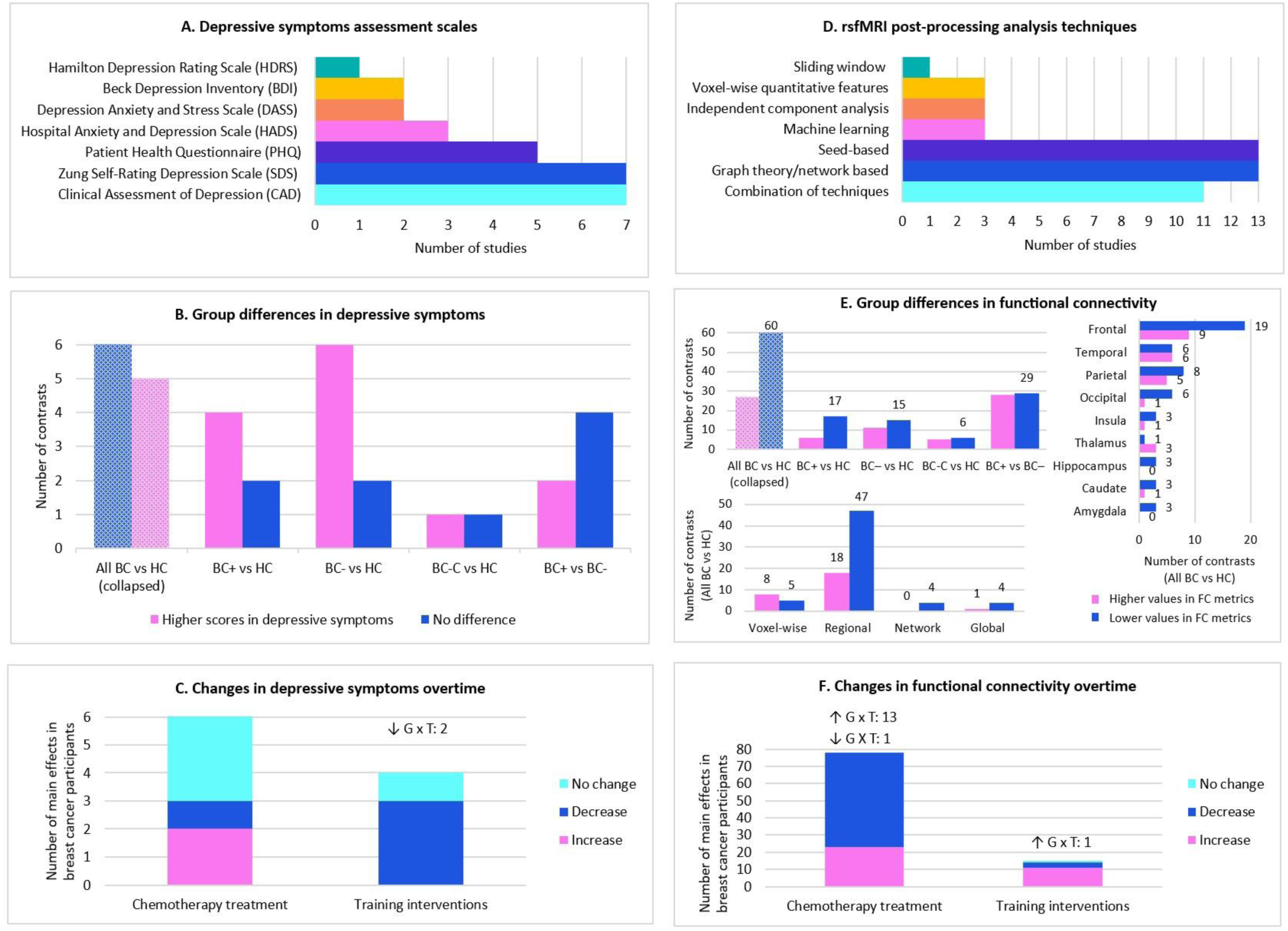
Summary of assessment methods, group differences, and changes over time for depressive symptoms and functional connectivity reported across included studies. *Note*. **Left panels (depressive symptoms)**: (A) Depressive symptoms assessment scales used across studies. (B) Cross-sectional group differences in depressive symptoms. Bars reflect the number of significant contrasts in which breast cancer groups reported higher scores (pink) or no difference (blue) compared with healthy controls or cancer control groups. The first bar (All BC vs HC) represents a collapsed category combining contrasts across BC+ (chemotherapy), BC- (no chemotherapy), and BC-C (combination treatment) subgroups versus healthy controls. (C) Within-group changes in depressive symptoms over time following chemotherapy or training interventions. Unique group-by-time (G × T) interactions are noted above bars. **Right panels (functional connectivity)**: (D) Resting-state fMRI analysis techniques used across studies. (E) Cross-sectional group differences in functional connectivity by group (top left), spatial scale (bottom left: voxel-wise, regional, network, global), and anatomical distribution of regional findings (right: cortical and subcortical). Bars reflect the number of significant contrasts showing higher values of functional connectivity metrics (pink), which broadly indicated increased connectivity, or lower values of functional connectivity metrics (blue), which broadly indicated decreased connectivity/dysconnectivity. (F) Within-group changes in functional connectivity over time following chemotherapy or training interventions. Unique G × T interactions are noted above bars.

The majority of studies (22/27, 81%) excluded participants (both people with BC and healthy controls) with current or historical severe psychiatric disorders (e.g., schizophrenia). Four studies specifically excluded those with self-reported major depression and four studies excluded participants that met a cut-off score for elevated depressive symptoms (e.g. scoring above 59 on the SDS [44]). In contrast, five studies [21,24,34–36] included participants with mild to severe depression based on cut-off scores from different scales. In three of these studies [24,35,36], depressive symptoms were confirmed through clinical assessment by a psychiatrist, and in one study [21], by a neurologist.

In cross-sectional studies, depressive symptoms were assessed using both qualitative and quantitative approaches. Three studies [21,35,36] reported the proportion of BC participants meeting criteria for clinically significant depressive symptoms, with prevalence rates ranging from 14.9% to 41%, as measured by the SDS and PHQ-9. Cross-sectional studies also showed that BC participants (newly diagnosed, undergoing active treatment, post-treatment) reported higher mean depressive symptom scores relative to healthy controls, with 11 significant group contrasts observed (Figure 3). Comparisons between BC participants who had received chemotherapy and those who had not showed mixed results, with only two contrasts indicating higher depressive symptoms in the chemotherapy group.

Longitudinal findings on depressive symptoms following cancer treatment were varied. Among studies tracking changes in depressive symptoms from baseline (post-diagnosis/pre-treatment) to post-treatment follow-up, two studies reported significant short-term increases: one study from baseline to one-week post-chemotherapy [34] and another study to one-month post-chemotherapy [38]. The same study [38] also reported decreases between one and seven-months post-chemotherapy. The remaining studies noted no significant changes [19,39,43,44]. Of the five longitudinal training intervention studies, two reported significant reductions in depressive symptoms: Kim et al. [34] in BC participants with mild to moderate MDD following a three-week mobile game program (‘Hit the Cancer’), and Van der Gucht et al. [41] in BC participants with cognitive impairment after an eight-week mindfulness-based program, with further improvement maintained at three-month follow-up. In contrast, Wolf et al. [42] reported no significant change in BC participants with cognitive impairment following a meta-cognitive training program, while two studies did not report on their findings [24,38].

### Functional connectivity outcomes

Acquisition parameters of the rsfMRI sequence of the studies are described in Table 1. Ten of 27 studies acquired rsfMRI within a larger multimodal MRI protocol (Table 1, ◊). All studies were conducted on 3T scanners. Acquisition time ranged from 6 to 12 min (mean = 8 min). The repetition time (TR) ranged between 870 and 3000 ms (mean = 1975 ms). The range of spatial resolution (i.e., voxel size) was between 1.86 mm^3^ and 4 mm^3^. Most studies acquired rsfMRI data with participants’ eyes closed (16/27 studies, 59%). Notably, many studies either did not report important scanning parameters or reported them inconsistently.

Table 1 and Figure 3 provide an overview of the analytic approaches and main FC results across studies. Five studies did not report statistical details of their rsfMRI findings [16, 25, 30, 35, 36]. The remaining studies utilised a range of techniques to analyse rsfMRI data. Region-of-interest (ROI) analyses were reported in 13 studies, where a seed region was selected *a priori* (most frequently the hippocampus, 4 studies), and used to compute FC maps from the temporal correlations between the ROI and all other brain regions. An equal number of studies (13) employed network/graph theoretical analyses, which examined the topology of functional brain networks at both global and regional levels, including whole-brain measures of integration and segregation (e.g., global efficiency, characteristic path length) and node-specific metrics of network topology (e.g., degree, nodal centrality). Three studies [16,35,36] applied machine learning techniques (e.g. multivariate pattern analysis, random forest classification) and leveraged complex algorithms to predict groups on the basis of FC patterns (e.g. high-risk versus low-risk depression profiles [35]). Additionally, three studies [19,21,27], employed independent component analysis (ICA), a data-driven method that decomposes the data from the entire brain into a number of components, with each component represented as a functional map. Three studies [22,24,40] extracted voxel-wise quantitative features from the rsfMRI data, such as the regional homogeneity (ReHo) and fractional amplitude of low-frequency fluctuations (ALFF). Only one study [32] used a sliding window analysis to assess dynamic functional connectivity, which captured temporal fluctuations in connectivity patterns overtime. Of note, eleven studies reported results from different analysis techniques within the same study.

As shown in Table 1 and Figure 3, cross-sectional rsfMRI findings demonstrated widespread FC alterations across comparisons between BC participants and healthy controls. Despite heterogeneous results, lower values of FC metrics were more frequently reported than higher values (60 vs 27), irrespective of analytic approach. In this review, lower values are interpreted as decreased connectivity (or dysconnectivity) within and between brain regions/networks in BC participants, while higher values reflect increased connectivity. Alterations were observed in both hemispheres, with a similar number of significant contrasts reported in the left (27 lower vs 8 higher) and right (23 lower vs 11 higher).

Across spatial scales, findings from the BC participants and healthy control comparisons were also varied. At the *voxel-wise level* (i.e., ALFF, ReHo), results were mixed (8 higher vs 5 lower). The strongest pattern emerged at the *regional level* (47 lower vs 18 higher; i.e., seed-based correlations, nodal graph-theory metrics, network-based statistics). Decreased connectivity was most frequently observed in the frontal lobe (19 lower vs 9 higher; e.g., medial prefrontal cortex, orbitofrontal gyrus, anterior cingulate). Alterations in the parietal lobe (8 lower vs 5 higher; e.g., posterior cingulate, angular gyrus, precuneus) and temporal lobe (6 lower vs 6 higher) were more mixed, while further alterations were noted in the occipital lobe (6 lower vs 1 higher) and the insula (3 lower vs 1 higher). In addition, subcortical regions showed a pattern of decreased connectivity, including the amygdala (3 lower), hippocampus (3 lower), and caudate (3 lower vs 1 higher). At the *network level* (i.e., a priori network-of-interest or ICA approaches), one study [29] reported decreased connectivity within the salience network, specifically dysconnectivity between the insula and anterior cingulate cortex. At the *global level* (whole-brain graph-theory metrics), findings also showed a trend of dysconnectivity (4 lower vs 1 higher). This included results of reduced clustering coefficient [14,28] and longer characteristic path length [28]. In contrast, one study reported increased global efficiency and longer path length [39].

Regarding longitudinal studies examining FC changes in BC participants pre- to post-cancer treatment (see Table 1 and Figure 3), findings across analysis techniques were similarly mixed but more often showed decreased connectivity following chemotherapy [19,30,39,43]. In contrast, training intervention studies generally reported increased connectivity at post-training intervention in BC participants, either relative to a waitlist control or compared with baseline in within-group designs [24,38,41], although one study reported no post-intervention change [42].

### Depressive symptoms-functional connectivity associations

Table 1 provides a summary on the associations between depressive symptoms and FC in people with BC. The majority of studies (23/27, 85%) conducted statistical correlation analyses (e.g. Pearson, Spearman correlation) to examine these associations. The remaining four studies (15%) employed alternative approaches: one study [21] compared FC between BC participants with and without mild depressive symptoms; two studies applied multivariate pattern analysis to classify high-versus low-risk depression trajectories [35] or predict major depressive disorder (MDD) 12 months post-diagnosis [36]; and one study [24] tracked short term FC changes within a BC cohort with moderate depression.

Most of the studies that conducted correlation analyses (16/23, 70%) reported non-significant associations. However, seven studies reported 11 significant correlations, which are visually presented in Supplementary Figure S1. As can be seen in this figure, the correlations varied in terms of direction (positive and negative), strength (r =.3 to .71; moderate to strong), and the implicated brain region/networks. Of these studies, only one study [22] was rated as good quality on the NOS, while six studies were rated as fair.

In cross-sectional studies reporting significant correlations, higher depressive symptom scores were linked with both decreased and increased connectivity. Specifically, higher PHQ or HADS scores were associated with increased ALFF in prefrontal and parietal regions [22,40] and increased seed-based connectivity between the thalamus and right postcentral gyrus [37]. In contrast, higher PHQ scores were associated with decreased ALFF across frontal, parietal, and occipital cortices, as well as in the subcortical left caudate [22].

Longitudinal studies similarly showed mixed correlations. One study [26] reported that higher SDS scores were associated with increased seed-based connectivity between the left hippocampus and left insula, and between the right posterior hippocampus and left superior temporal gyrus from pre- to one-week post-chemotherapy.

Another study [19] reported that higher SDS scores were associated with decreased ICA-based connectivity between the self-referential and visual networks from one week to six months post-chemotherapy. Correlations from training intervention studies also varied. In one trial [34], lower BDI scores were associated with decreased seed-based connectivity between the posterior cingulate cortex and right anterior insula following a mobile game intervention, whereas in another study [41], lower DASS scores were linked to increased seed-based connectivity between the anterior cingulate cortex and intraparietal sulcus following mindfulness training.

Among studies not using correlation analyses, depression-related FC alterations were also observed in BC participants. For example, Bukkieva et al. [21] reported that BC participants with mild depression (vs without) showed decreased seed-based connectivity between the medial prefrontal cortex and the cuneal cortex, planum polare, and fusiform gyrus, as well as altered connectivity between the medial prefrontal cortex and both the parahippocampal region and the dorsal attention network. Liang et al. [35] employed multivariate pattern analysis and found that FC patterns in the frontal medial cortex and frontal pole distinguished high-versus low-risk depression trajectories among BC participants.

## Discussion

This is the first systematic review to date to synthesise resting-state functional magnetic resonance imaging (rsfMRI) studies investigating the relationship between depressive symptoms and functional connectivity (FC) in people with breast cancer (BC). Among cross-sectional studies, BC participants demonstrated elevated depressive symptoms and widespread FC alterations, most often reflecting patterns of dysconnectivity, compared to healthy controls. However, there was no strong evidence for associations between depressive symptoms and FC. Longitudinal studies revealed dynamic patterns over time, with some showing ongoing depressive symptoms and altered FC, while others suggested recovery-related changes in brain function.

### Elevated depressive symptoms experienced by people with breast cancer: the need for training interventions

The present review highlights the higher levels of self-reported depressive symptoms experienced by people with BC compared with healthy controls. This finding is consistent with a previous systematic review [49] that found significantly elevated depressive symptoms in BC participants versus healthy controls in 19 of 38 studies. Cross-sectional results from the reviewed studies also showed elevated depressive symptoms across diverse BC groups, irrespective of cancer treatment type or stage of care, with no clear differences observed between those who did and did not receive chemotherapy. This contrasts with previous literature which has commonly emphasised chemotherapy-related effects on emotional and cognitive wellbeing [50]. Specifically, anthracycline- and taxane-based regimens, have been linked to elevated depressive symptoms, potentially through neurotoxic and inflammatory mechanisms [51]. For example, Tzang et al. [52] reported higher depressive symptom severity in chemotherapy-treated BC participants compared to cancer controls and healthy controls, with symptoms correlating with elevated proinflammatory cytokines (e.g., interleukin-6) in the chemotherapy group only.

However, no chemotherapy-specific effects were observed in the present review. This may reflect the fact that many BC participants who did not receive chemotherapy underwent other treatments (e.g. surgery, radiation, hormonal therapy) which have also been linked to biological disruption and psychological distress, potentially contributing to similarly elevated symptom profiles across groups [3]. Hence, it is suggested that depressive symptoms in people with BC are not unidimensional or attributable to a single factor but rather reflect a multidimensional outcome of broader disease- and treatment-related processes [53]. This aligns with biopsychosocial models, such as proposed by Ahles and Root [54], which conceptualise cancer-related cognitive and mood disturbances as the cumulative result of biological (e.g., inflammation, genetic susceptibility), psychological (e.g., coping, appraisal), and social (e.g., limited social support, life stressors) deficits.

The synthesis of longitudinal studies showed that elevated depressive symptoms often persist well beyond the acute treatment phase, with many people with BC demonstrating minimal or no improvement several months to a year following diagnosis and/or treatment. This aligns with previous literature highlighting that emotional distress can be long lasting in a subset of patients [12,55]. For example, an earlier systematic review of 17 studies (including five longitudinal) estimated that 20-30% of people with BC experience persistent depressive symptoms for up to five years post-diagnosis [12]. Moreover, a large longitudinal study [55] (n = 653) using the Center for Epidemiological Studies Depression Scale (CES-D) found that while most BC participants reported low or borderline symptoms over the 18 months following diagnosis, approximately 20% exhibited clinically significant depressive symptoms that persisted throughout this period. These longitudinal patterns challenge assumptions of natural remission and underscore that, for some people with BC, depressive symptoms can be enduring and may not improve without targeted intervention.

Numerous psychosocial and training interventions (i.e., cognitive behavioural therapy, mindfulness/relaxation, physical activity, support groups, digital tools) have been trialled to alleviate depressive symptoms in BC populations [56]. In this review, two randomised controlled training interventions reported significant improvements [34,41]. A 30-day mobile attention-training game yielded moderate-to-large-effects in BC participants with mild-to-moderate depression, with the authors suggesting that repeated practice in shifting attention away from cancer-related threat cues and towards positive stimuli helped reduce rumination and threat-focused thinking [34]. Similarly, an adapted eight-week Mindfulness-Based Stress Reduction program, incorporating practices such as body scan, mindful movement, and breathing exercises, observed large effects at three-month follow-up in BC participants with cognitive impairment, proposed to reflect improved emotion regulation and increased acceptance-based coping [41]. Although effect sizes in these trials exceeded those reported in previous meta-analyses of psychosocial interventions for BC [56], this may partly reflect the smaller sample sizes common in neuroimaging research. Nonetheless, these findings highlight the potential of targeted interventions and point to the need for more rigorous trials in BC populations. To advance the field, interventions may benefit from addressing unhelpful cognitive-emotional processes and behaviours (e.g., cognitive distortions, emotional suppression, avoidance) while strengthening supportive psychological resources (e.g., coping skills, self-compassion), which could buffer against depressive symptoms during breast cancer survivorship. For example, a recent mindfulness-based intervention showed that reductions in rumination and intrusive thoughts, together with improvements in self-compassion, purpose, and meaning, mediated improvements in depressive symptoms among younger BC participants over six months [57]. Future work should assess the durability of these benefits (e.g., >6 months) and clarify mechanisms of change to inform scalable interventions for survivorship care.

Besides training intervention studies, further research should also try to capture the fluctuating course of depressive symptoms frequently reported by people with BC [58,59]. All of the studies in this review utilised a self-report questionnaire administered at a single time point, which only captures a limited snapshot of symptomatology. These measures are also prone to recall bias, as they rely on participants’ ability to accurately reflect on their past emotional state, such as over the last one to two weeks [60]. Moreover, collapsing a participants experience into one overall score can mask meaningful intra-individual variability and context-dependent fluctuations, limiting ecological validity [58]. Future studies should conduct ecological momentary assessment (EMA) of depressive symptoms to capture dynamic, within-person changes over time and better reflect individuals’ everyday experiences. Previous longitudinal EMA studies conducted on people with depression have demonstrated that daily mood/depressive symptom ratings are sensitive to symptom fluctuations and correspond with both weekly self-report and clinician-rated depression severity [61,62]. Recently, a novel app (*Mind Trax*) that assessed daily cognitive functioning and emotional wellbeing over 30 days demonstrated good feasibility, usability and construct validity in both people with breast cancer and healthy controls [63]. Hence, utilising EMA to capture these temporal patterns often missed by single time-point assessments, may help inform the delivery of more timely and personalised treatment.

### Alterations in functional connectivity in people with breast cancer across diverse studies using a wide array of analysis techniques

Although cross-sectional findings were heterogenous, the present review identified that people with BC more frequently showed lower values in FC metrics (decreased connectivity) compared with healthy controls, across different spatial scales and analytic approaches. In other words, while some studies reported higher values (increased connectivity), the more common pattern was lower values, reflecting dysconnectivity within and between brain regions/networks. At the regional level, decreased connectivity was most frequently observed in frontal lobe regions, including the medial prefrontal cortex, orbitofrontal cortex, and anterior cingulate cortex, which are broadly involved in self-referential thinking, emotion regulation, and reward-based decision-making, respectively [64]. These regions also serve as central hubs within large-scale brain networks implicated in cognitive-affective integration (e.g., the default mode network, discussed in more detail below) [66,67]. This finding of frontal dysconnectivity aligns with prior systematic reviews of multimodal neuroimaging studies in cancer populations [11,65], which have repeatedly documented the frontal lobe as a primary site of cancer-related brain alterations. For example, structural MRI studies have reported cortical thinning in the medial and superior frontal cortices [68], diffusion tensor imaging has revealed reduced integrity of frontostriatal white matter tracts [69], and task-based fMRI studies have demonstrated decreased activation in the dorsolateral prefrontal cortex and anterior cingulate cortex during executive function tasks in BC participants compared to healthy controls [70]. This vulnerability may reflect the frontal region’s high metabolic demands and extensive network integration, which may render it particularly susceptible to disease and treatment-related mechanisms, including inflammation, oxidative damage, and neurotoxic effects [68]. Animal studies further support this; for example, chemotherapy-treated tumor-bearing mice, compared to non-exposed controls, exhibited increased proinflammatory signaling and altered expression of genes involved in synaptic plasticity in the prefrontal cortex [71,72]. Although less frequently reported, this review also noted dysconnectivity in subcortical regions (i.e., hippocampus, amygdala, caudate) as well as both increased and decreased connectivity in the parietal lobe (i.e., posterior cingulate cortex, angular gyrus, and precuneus). These findings suggest that FC alterations in BC populations extends beyond focal regions and may reflect broader disruptions in large-scale brain networks.

Across studies, many of the regions where decreased connectivity was observed in BC participants relative to healthy controls, overlapped with key hubs of large-scale brain networks, most notably, the default mode network (DMN). The DMN - comprising the precuneus, posterior cingulate cortex, medial frontal cortex, middle temporal gyrus, lateral parietal regions, and hippocampus - supports internally directed processes such as autobiographical memory retrieval, self-referential thinking, and affective regulation [67]. Dysconnectivity in DMN-associated regions is consistent with prior reviews proposing DMN dysfunction as a potential neural correlate of cognitive and affective difficulties in cancer patients [65,73]. For example, a previous activation likelihood estimation meta-analysis of task-based fMRI studies in non-central nervous system cancer populations identified a pattern of decreased activation in the precuneus/parietal lobe (a key hub of the DMN) during cognitive tasks [90]. In addition to the DMN, FC alterations were also reported in regions implicated in the salience network (SN; e.g., anterior insula, inferior frontal gyrus) and dorsal attention network (DAN; e.g., intraparietal sulcus, superior parietal lobule). Together, these three systems form the brain’s core “triple-network”, a model proposed by Menon [66], which highlights their central role in coordinating the dynamic balance between internally focused thought (DMN), externally directed attention (DAN), and the detection of salient stimuli (SN). An increasing number of studies have shown that dysfunction within the triple network is implicated with cognitive and affective impairments across a range of clinical populations, including MDD, traumatic brain injury, and schizophrenia [74–76]. Findings from this review suggest that similar large-scale network alterations may also underlie cognitive-affective symptoms in BC populations.

The present review further found evidence of alterations in the global organizational architecture of the brain in people with BC compared to healthy controls. Reductions in clustering coefficient [14,28] and longer characteristic path length [28] point to weaker local specialization and less efficient communication across brain regions [77]. In contrast, another study reported higher global efficiency alongside longer path length [39], reflecting a divergent pattern of altered integration in which measures of increased information transfer and reduced efficiency co-occur [77]. Although based on limited studies, these findings suggest that breast cancer and its treatments may affect the connectome’s topological organization, raising the possibility of vulnerabilities in how the brain balances segregation (specialized processing within local networks) and integration (communication across distributed networks) at a global scale. As with network-level results, alterations in global brain connectivity have also been reported across a range of psychiatric and neurological disorders [77].

### The missing link between depressive symptoms and functional connectivity alterations in people with breast cancer

Examining brain-behavior relationships is essential for understanding how FC alterations relate to depressive symptoms in people with BC. However, despite elevated depressive symptoms and widespread FC alterations shown across studies, the present review found limited evidence for a strong link between these metrics. Only a minority of studies (7/27, 26%) reported significant correlations, and these varied in terms of direction, magnitude, and implicated regions/networks.

Among this small numbers of studies, depressive symptoms-functional connectivity relationships in people with BC can be broadly interpreted as reflecting three different mechanistic patterns. *Decreased connectivity* was associated with higher symptom severity in two studies [19,22], which may reflect *direct neural injury or damage* from disease- or treatment-related processes (e.g., chemotherapy-induced neurotoxicity, chronic inflammation) [50,69]. Specifically, elevated depressive symptoms were linked with dysconnectivity between self-referential and visual networks (longitudinally one-week to six-months after chemotherapy) [19]) and across frontal, parietal, and occipital cortices, as well as the caudate nucleus [22]. These weakened connections may undermine circuits supporting affect regulation, attentional control, and disengagement from negative thought patterns, thereby possibly contributing to elevated depressive symptom [74,78]. In addition, *increased connectivity* was associated with higher symptom severity in four studies [22,26,37,40], which may indicate *maladaptive reorganisation* whereby regions or networks become overly synchronized in rigid or ineffective patterns [79]. For example, elevated depressive symptoms were associated with heightened synchrony between the thalamus and precentral gyrus [37], and between the hippocampus and insula/temporal regions shortly after chemotherapy [26]. Although these increases may initially reflect adaptive compensatory attempts to stabilise disrupted circuits, persistent hyperconnectivity can become metabolically costly and reinforce inflexible pathways that impair emotion regulation and cognitive flexibility, thereby potentially contributing to heightened depressive symptoms [74,78,79]. Finally, potential *adaptive reorganisation* was observed in two training intervention studies [42,49], where symptom improvements were associated with decreased connectivity between posterior cingulate and insula regions, and increased connectivity between the anterior cingulate and intraparietal sulcus. These findings suggest that therapeutic benefit may possibly occur either through pruning inefficient connections or strengthening control and attentional networks, thereby supporting the brain’s capacity for adaptive neuroplasticity in recovery [79].

However, the current rsfMRI literature is too mixed to establish reliable neural/functional correlates in BC populations. This is in contrast to previous neuroimaging studies in MDD [74,78], where robust associations have been consistently demonstrated, including between elevated depressive symptoms and decreased connectivity between the prefrontal cortex (particularly dorsolateral and medial regions) and limbic structures such as the amygdala and hippocampus, alongside hyperconnectivity within limbic circuits (e.g., amygdala hypercoupling with ventral and subcortical regions). These patterns have been frequently linked to impaired top-down regulation of emotion, increased negative affect, rumination, and memory biases [78].

Non-significant associations across this reviewed literature are suggested to be due to the nature of the analysis. Most studies assessed the relationship between depressive symptoms and FC alterations using bivariate correlations between a single depression score and a single brain metric. While intuitive, this approach oversimplifies the dynamic and multidimensional nature of brain-behavior relationships [80]. As aforementioned, depressive symptoms in people with BC likely arise from complex interactions between neurobiological mechanisms and psychological deficits [53,54], which cannot be fully captured through univariate analyses [80]. Future research could benefit from using multilayer network analysis (MLN), which enables simultaneous modeling across levels of organization (i.e., multiple behavior/symptoms and brain variables) and/or across time (e.g., days, weeks using EMA) [81]. In this approach, separate layers (with their own single-layer nodes and edges, or variables and correlations) can be connected into a multilayer network by interlayer edges [81]. Specifically, this MLN approach is a new way to study the neural correlates that couple one symptom variable to another (e.g., neural mechanisms that link depressive symptoms to fatigue).

The application of multilayer networks is still in its infancy. To date, only three other studies [82–84] have utilised this approach, including one in depression. Hilland et al. [82] applied an MLN framework to examine associations between self-reported depressive symptoms (via the Beck Depression Inventory) and regional brain volumes in a group of healthy adults with varying levels of depressive symptoms. The author’s analysis identified cross-layer links between depressive symptoms such as sadness, crying, and irritability, and structural features of the hippocampus, insula, cingulate cortex, and fusiform gyrus. These findings highlight the potential of MLN methods to capture symptom-specific neural correlates that may be overlooked by univariate models.

Besides the usage of multilayer network analysis, future work should adopt standardized pipelines, such as the HALFpipe (Harmonized Analysis of Functional MRI pipeline) [85]. This pipeline provides standardized protocols that streamline data processing, increase data fidelity, harmonize data across multiple sites, and support the use of different complementary analytic methods (e.g., seed-based, graph-theoretical) on the same dataset. This will increase the transparency and reproducibility of rsfMRI findings (for a review, see [86]). For example, the ENIGMA Obsessive Compulsive Disorder (OCD) study used this toolbox across 28 sites (1024 OCD patients vs 1028 controls) and showed widespread FC disruptions, particularly global hypoconnectivity in sensorimotor networks [87]. Importantly, an ENIGMA-MDD resting-state project is currently underway, explicitly using HALFpipe to examine connectivity differences between individuals with MDD and healthy controls [88].

### Clinical implications

This review highlights the need for timely and ongoing psychosocial support for people with breast cancer throughout survivorship, including after completing treatment. In line with psycho-oncology guidelines [3], clinicians are encouraged to monitor depressive symptoms across the care trajectory and refer patients to training programs or individual psychological intervention when appropriate. Importantly, many of the measures used to assess depressive symptoms in this literature had not been validated among people with cancer (e.g. the Clinical Assessment of Depression), and often combined affective, cognitive, and somatic experiences into a single score. As a result, depressive symptoms may be confounded with other treatment-related disturbances such as fatigue, pain, or sleep issues, which can lead to either overestimation or underestimation of emotional distress. The Patient-Reported Outcomes Measurement Information System (PROMIS) framework [89], validated in cancer populations, offers a more robust approach by enabling depressive symptoms to be assessed separately from somatic domains. The Depression Item Bank focuses specifically on affective and cognitive features (e.g., sadness, hopelessness, anhedonia), while fatigue, pain, and cognitive function can be measured in parallel using distinct banks, thereby helping to reduce diagnostic confounding and supporting more accurate, tailored psychosocial care planning. Finally, when translating neuroimaging findings into practice, clinicians should note that most rsfMRI studies excluded individuals with severe psychiatric conditions, such as MDD. Consequently, the symptom profiles reported may underestimate the severity and complexity typically seen in oncology settings.

### Limitations and conclusion

Several limitations of this systematic review should be acknowledged. First, the small number of eligible studies necessitated broad inclusion criteria, introducing heterogeneity in participant characteristics, depressive symptom measures, and rsfMRI analytic techniques. This variability limited comparability across studies and precluded a meta-analysis. Second, depressive symptoms were often examined as a secondary outcome in large-scale studies primarily focused on other clinical domains (most commonly cognitive functioning) which may have reduced the depth and consistency of depression-specific neuroimaging investigations. Third, a large number of rsfMRI studies (*n* = 45) were excluded from this review because they did not assess associations between depressive symptoms and functional connectivity. As a result, the findings presented here may not reflect the full extent of functional connectivity alterations reported in the broader breast cancer literature. Future reviews could extend this work by incorporating studies outside the current scope and synthesising connectivity patterns across additional symptom domains.

Despite these limitations, this review highlights the importance of investigating depressive symptoms-functional connectivity relationships via resting-state fMRI to advance understanding in people with breast cancer. Although elevated depressive symptoms and diffuse FC alterations (predominantly patterns of dysconnectivity) were frequently reported, associations between the two remain unclear. Future studies that integrate ecological momentary assessment, adopt standardized rsfMRI pipelines, and apply network-based analytic approaches are essential for unravelling brain-symptom relationships in this population and unlocking the potential of resting-state fMRI to identify reliable neural correlates of depressive symptoms. Pursuing these directions will ultimately inform more personalized care during cancer survivorship and enhance long-term patient outcomes.

## Supporting information

Supplementary Figure S1 Correlations

Supplementary Table S1 PRISMA Checklist

Supplementary Table S2 Search Strategies

Supplementary Table S3 NOS Quality Assessments

Supplementary Table S4 Detailed Results Summaries

Supplementary Table S5 Clinical Variables

## Data Availability

All data extracted and qualitatively analysed during this study are available from the corresponding author upon request.

## Acknowledgements

Author Jacqui Saward is supported by a Deakin University Post-Graduate Research Scholarship

## Author contributions

Jacqueline Saward, Juan Dominguez, and Karen Caeyenberghs contributed to the conceptualisation of the study design, screening, data extraction, interpretation of findings, and drafting and critical revision of the manuscript. Annalee Cobden assisted with screening and study selection and provided critical revisions. Priscilla Gates contributed to the interpretation of results, clinical implications, and manuscript revisions. Jacqueline Saward prepared the first draft of the manuscript. All authors reviewed and approved the final version of the manuscript.

## Ethics approval

Not applicable

## Consent to participant/consent for publication

Not applicable

