## Supplementary Figure S1 Correlations for "Investigating the link between depressive symptoms and resting-state brain connectivity in people with breast cancer: A systematic review"

**Supplementary Figure S4.** Resting-state fMRI studies reporting significant correlations between depressive symptoms and functional connectivity in people with breast cancer

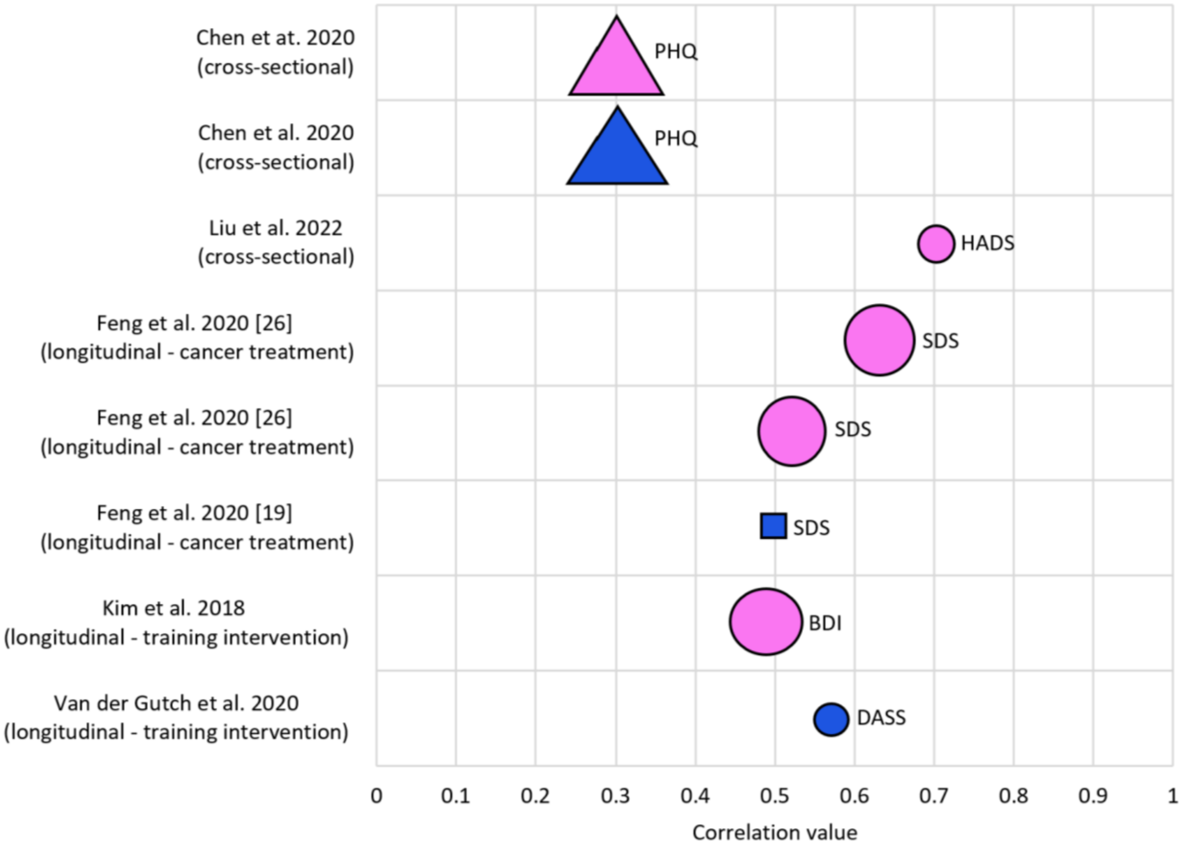


*Note.* Positive correlations are shown in pink, negative correlations in blue. Depressive symptom scales are noted beside each marker: PHQ = Patient Health Questionnaire, HADS = Health Anxiety and Depression Scale, SDS = Zung Self-Rating Depression Scale, BDI = Beck Depression Inventory, DASS = Depression Anxiety and Stress Scale. Marker shapes represent rsfMRI analysis techniques: triangle = amplitude of low-frequency fluctuations, circle = seed-based analysis, and square = independent component analysis. Marker size indicates study sample size: small (15-20), medium (20-30), large (30-45).
