## Supplementary Table S2 Search Strategies for "Investigating the link between depressive symptoms and resting-state brain connectivity in people with breast cancer: A systematic review"

**Supplementary Table S1.** Full search strategies for each database

| Database | Search Syntax |
|---|---|
| **Embase** | S1 AND S2 AND S3 |
| S1 | (‘breast cancer’:ti OR ‘breast oncology’:ti OR ‘breast neoplasm*’:ti OR ‘breast carcinoma*’:ti OR ‘breast tumor*’:ti OR ‘breast tumour*’:ti OR ‘breast malignancy’:ti OR ‘breast cancer’:ab OR ‘breast oncology’:ab OR ‘breast neoplasm*’:ab OR ‘breast carcinoma*’:ab OR ‘breast tumor*’:ab OR ‘breast tumour*’:ab OR ‘breast NEAR/3 cancer’ OR ‘breast malignancy’/de) |
| S2 | (‘resting state’:ti OR ‘rsfMRI’:ti OR ‘functional connectivity’:ti OR ‘functional network’:ti OR ‘graph theory’:ti OR ‘connectome’:ti OR ‘resting state’:ab OR ‘rsfMRI’:ab OR ‘functional connectivity’:ab OR ‘functional network’:ab OR ‘graph theory’:ab OR ‘connectome’/de) |
| S3 | [embase]/lim |
| **PsycINFO** | SI AND S2 |
| S1 | (TI ‘breast cancer’ OR TI ‘breast oncology’ OR TI ‘breast neoplasm*’ OR TI ‘breast carcinoma*’ OR TI ‘breast tumor*’ OR TI ‘breast tumour*’ OR TI ‘breast malignancy’ OR AB ‘breast cancer’ OR AB ‘breast oncology’ OR AB ‘breast neoplasm*’ OR AB ‘breast carcinoma*’ OR AB ‘breast tumor*’ OR AB ‘breast tumour*’ OR AB ‘breast malignancy’ OR ‘breast N3 cancer’ OR MA ‘breast cancer’) |
| S2 | (TI ‘resting state’ OR TI ‘resting-state’ OR TI ‘rs-fMRI’ OR TI ‘rsfMRI’ OR TI ‘functional connectivity’ OR TI ‘functional network’ OR TI ‘graph theory’ or TI ‘connectome’ OR AB ‘resting state’ OR AB ‘resting-state’ OR AB ‘rs-fMRI’ OR AB ‘rsfMRI’ OR AB ‘functional connectivity’ OR AB ‘functional network’ OR AB ‘graph theory’ OR AB ‘connectome’ OR MA ‘functional connectivity’) |

| | |
|---|---|
| **Medline Complete** | S1 AND S2 |
| S1 | (TI ‘breast cancer’ OR TI ‘breast oncology’ OR TI ‘breast neoplasm*’ OR TI ‘breast carcinoma*’ OR TI ‘breast tumor*’ OR TI ‘breast tumour*’ OR TI ‘breast malignancy’ OR AB ‘breast cancer’ OR AB ‘breast oncology’ or AB ‘breast neoplasm*’ OR AB ‘breast carcinoma*’ OR AB ‘breast tumor*’ OR AB ‘breast tumour*’ OR AB ‘breast malignancy’ OR ‘breast N3 cancer’ OR MH ‘breast cancer’) |
| S2 | (TI ‘resting state’ OR TI ‘resting-state’ OR TI ‘rs-fMRI’ OR TI ‘rsfMRI’ OR TI ‘functional connectivity’ OR TI ‘functional network’ OR TI ‘graph theory’ or TI ‘connectome’ OR AB ‘resting state’ OR AB ‘resting-state’ OR AB ‘rs-fMRI’ OR AB ‘rsfMRI’ OR AB ‘functional connectivity’ OR AB ‘functional network’ OR AB ‘graph theory’ OR AB ‘connectome’ OR MH ‘functional connectivity’) |
| **CINAHL** | S1 AND S2 |
| S1 | (TI ‘breast cancer’ OR TI ‘breast oncology’ OR TI ‘breast neoplasm*’ OR TI ‘breast carcinoma*’ OR TI ‘breast tumor*’ OR TI ‘breast tumour*’ OR TI ‘breast malignancy’ OR AB ‘breast cancer’ OR AB ‘breast oncology’ OR AB ‘breast neoplasm*’ OR AB ‘breast carcinoma*’ OR AB ‘breast tumor*’ OR AB ‘breast tumour*’ OR AB ‘breast malignancy’ OR ‘breast N3 cancer’ OR MH ‘breast cancer’) |
| S2 | (TI ‘resting state’ OR TI ‘resting-state’ OR TI ‘rs-fMRI’ OR TI ‘rsfMRI’ OR TI ‘functional connectivity’ OR TI ‘functional network’ OR TI ‘graph theory’ OR TI ‘connectome’ OR AB ‘resting state’ OR AB ‘resting-state’ OR AB ‘rs-fMRI’ OR AB ‘rsfMRI’ OR AB ‘functional connectivity’ OR AB ‘functional network’ OR AB ‘graph theory’ OR AB ‘connectome’ OR MH ‘functional connectivity’) |

*Note.* This table presents the full search strings used to retrieve resting-state fMRI studies on people with breast cancer across the four databases (EMBASE, PsycINFO, Medline, and CINAHL), with the last search conducted on August 13, 2025.
