## Supplementary Table S3 NOS Quality Assessments for "Investigating the link between depressive symptoms and resting-state brain connectivity in people with breast cancer: A systematic review"

**Supplementary Table S3a.** Newcastle-Ottawa Scale (NOS) quality assessment adapted for cross-sectional studies

| | **Selection** | | | | **Comparability** | **Outcome** | | **Score** | **Study quality** |
|---|---|---|---|---|---|---|---|---|---|
| Author, year | Representativeness of the sample | Sample size | Non-respondents | Ascertainment of exposure | Confounding controlled | Outcome assessment | Statistics | Total | AHRQ rating (risk of bias) |
| Bruno et al. 2012 | * | | * | | * | * | * | 5/9 | Fair (Moderate) |
| Bukkieva et al. 2022 | | | * | ** | * | * | | 5/9 | Fair (Moderate) |
| Chen et al. 2017 | | | * | * | ** | * | * | 6/9 | Fair (Moderate) |
| Chen et al. 2020 | | * | * | ** | ** | * | * | 8/9 | Good (Low) |
| Donofry et al. 2022 | | | * | ** | ** | * | * | 7/9 | Good (Low) |
| Hampson et al. 2015 | * | | * | * | ** | * | * | 7/9 | Good (Low) |
| Harrison et al. 2021 | * | | | | * | * | * | 4/9 | Poor (High) |
| Henneghan et al. 2022 | | | * | | ** | * | * | 5/9 | Fair (Moderate) |
| Kesler et al. 2013 | * | | * | | ** | * | * | 6/9 | Fair (Moderate) |
| Kesler et al. 2017 | | | * | | ** | * | * | 5/9 | Fair (Moderate) |
| Kesler et al. 2021 | | | * | | * | * | * | 4/9 | Poor (High) |
| Liang et al. 2024 [35] | * | | * | | | * | | 3/9 | Poor (High) |
| Liu et al. 2022 | | | * | * | ** | * | * | 6/9 | Fair (Moderate) |
| Shen et al. 2021 | | | | | ** | * | | 3/9 | Poor (High) |
| Zhou et al. 2022 | | | * | ** | * | * | * | 6/9 | Fair (Moderate) |

*Note:* The NOS adapted for cross-sectional studies was used to assess the quality of 15 cross-sectional studies in this review. Criteria for each subscale were identified based on the review's aims and the specific sample and methodological characteristics of the included rsfMRI studies. Studies were awarded 0 stars if the criteria were not met, 1 star if the criteria were met, and, in some cases, 2 stars if the criteria were met using a validated method or an established model. The total score across all subscales was used to classify overall study quality according to the Agency for Healthcare Research and Quality (AHRQ) standards: good/low risk of bias (>7), fair/moderate risk of bias (5–7), and poor/high risk of bias (<5). Details of the criteria for each subscale item are provided below:

- *Representativeness of the sample:* 1 star = truly representative of the average breast cancer participant (all subjects or random sampling), 1 star = somewhat representative of the average breast cancer participant (non-random sampling e.g. selected group from hospitals, clinics or support groups), 0 stars = selected group from a single institution or no description of the derivation of the cohort
- *Sample size:* 1 star = justified and satisfactory (including sample size classification), 0 stars = not justified or no information provided
- *Non-respondents/missing data:* 1 star = reported completion rate > 80% (i.e., the number of participants who completed the depressive symptoms measure and resting-state fMRI scan), 0 stars = completion rate not reported or unclear regarding the number of participants who completed the depression symptoms measure and resting-state fMRI scan
- *Ascertainment of exposure:* 2 stars = confirmed breast cancer diagnosis and treatments through secure records (e.g. hospital records, pathology results), 1 star = structured interview or questionnaire, 0 stars = self-report only or unclear description
- *Confounding controlled:* 2 stars = study controls for age AND any additional relevant factors (e.g. education, menopausal status), 1 star = study controls for age OR additional relevant factor only, 0 stars = study does not control for confounding factors
- *Outcome assessment:* 1 star = used a validated assessment tool to measure depressive symptoms, 0 stars = Did not use a validated assessment tool to measure depressive symptoms
- *Statistics:*1 star = statistical tests used were clearly described, appropriate, and where relevant included measures of association with probability levels (p-values), 0 star = statical tests not appropriate, not described, or incomplete.

**Supplementary Table S3b.** Newcastle-Ottawa Scale (NOS) quality assessment for longitudinal studies

| | Selection | | | | Comparability | Outcome | | | Score | Study quality |
|---|---|---|---|---|---|---|---|---|---|---|
| Author, year | Representativeness of exposed cohort | Selection of non-exposed cohort | Ascertainment of exposure | Demonstration that outcome of interest was not present at start of study | Confounding controlled | Ascertainment of outcome | Was follow-up long enough for outcome to occur | Adequacy of follow up cohorts | Total | AHRQ rating (risk of bias) |
| **Pre-post cancer treatment** | | | | | | | | | | |
| Feng et al. 2020 [26] | | | | * | ** | * | * | * | 6/9 | Fair (Moderate) |
| Feng et al. 2020 [19] | | | | * | ** | * | * | | 5/9 | Fair (Moderate) |
| Kardan et al. 2019 | | * | | | * | * | * | * | 5/9 | Fair (Moderate) |
| Liang et al. 2024 [36] | * | NA | | | ** | * | * | * | 6/9 | Fair (Moderate) |
| Phillips et al. 2022 | | | | | ** | * | * | | 4/9 | Poor (High) |
| Yang et al. 2025 | | | | | ** | * | * | * | 5/9 | Fair (Moderate) |
| Zhuang et al. 2024 | | | | * | ** | * | * | * | 6/9 | Fair (Moderate) |
| **Pre-post training intervention** | | | | | | | | | | |
| Choghazardi et al. 2025 | | NA | * | * | NA | * | | | 3/9 | Poor (High) |

| | | | | | | | | | | |
|---|---|---|---|---|---|---|---|---|---|---|
| Kim et al. 2018 | | * | * | * | | * | * | * | 6/9 | Fair (Moderate) |
| Melis et al. 2023 | * | * | * | | ** | * | * | | 7/9 | Good (Low) |
| Van der Gutch et al. 2020 | | * | * | * | | * | * | | 5/9 | Fair (Moderate) |
| Wolf et al. 2016 | | NA | * | * | NA | * | * | * | 5/9 | Fair (Moderate) |

*Note:* The NOS for cohort studies was used to assess the quality of 12 longitudinal studies in this review. Although some of the subscale criteria differed from those for cross-sectional studies, the overall approach remained consistent. As outlined above, subscale criteria were based on the review's research aims and the sample and methodological characteristics of the included rsfMRI studies. Studies were awarded 0 stars if criteria were not met, 1 star if met, and 2 stars if met using validated methods or established models. The total score across subscales classified study quality based on AHRQ standards: good/low risk of bias (>7), fair/moderate risk of bias (5–7), and poor/high risk of bias (<5). Detailed criteria for each subscale item are provided below:

- *Representativeness of exposed cohort:* 1 star = truly representative of the average breast cancer participant (all subjects or random sampling), 1 star = somewhat representative of the average breast cancer participant (non-random sampling e.g. selected group from hospitals, clinics or support groups), 0 stars = selected group from a single institution or no description of the derivation of the cohort
- *Selection of non-exposed cohort:* 1 star = drawn from same population as the exposed cohort (i.e., comparable population without exposure), 0 stars = drawn from a different source or no description of the derivation of cohort provided
- *Ascertainment of exposure (for pre and post cancer treatment studies):* 1 star = confirmed breast cancer diagnosis/chemotherapy treatment through secure record (i.e., hospital records, pathology results), 1 star = structured interview of questionnaire, 0 stars = self-report or unclear description

  *Ascertainment of exposure (for training intervention studies):* 1 star = confirmed participation in the intervention through session attendance records, interview, or monitoring tools, 0 stars = no description of how intervention exposure was confirmed
- *Demonstration that outcome of interest was not present at start of study (for pre and post cancer treatment studies):* 1 star = participants did not exhibit the outcome of interest at baseline (e.g., elevated depression levels beyond typical or normative ranges, altered functional connectivity). If participants with elevated depressive

symptoms were included for research aims, they did not show alterations in functional connectivity at baseline or significantly differ from the control condition. 0 stars = no clear demonstration that participants were free from the outcomes of interest at baseline

- *Demonstration that outcome of interest was not present at start of study (for pre and post training intervention studies):* 1 star = the outcome of interest would be symptom improvement or reduction rather than absence at baseline. Baseline severity of symptoms was established to track changes over time and effect of intervention. 0 stars = no baseline assessment conducted
- *Confounding controlled:* 2 stars = study controls for age AND any additional relevant factors (e.g. education, menopausal status), 1 star = study controls for age OR additional relevant factor only, 0 stars = study does not control for confounding factors
- *Ascertainment of outcome:* 1 star = used a validated assessment tool to measure depressive symptoms, 0 stars = Did not use a validated assessment tool to measure depressive symptoms
- *Was follow up long enough for outcomes to occur:* 1 star = yes, 0 stars = no
- *Adequacy of follow up cohorts:* 1 star = complete follow-up of all participants accounted for, or participants lost to follow up unlikely to introduce bias (i.e., number lost less than or equal to 20% or description of those lost suggested no different from those followed), 0 stars = follow up less than 80% without sufficient description of those lost
