## Supplementary Table S4 Detailed Results Summaries for "Investigating the link between depressive symptoms and resting-state brain connectivity in people with breast cancer: A systematic review"

**Supplementary Table S4.** Detailed summary of study selection, participant characteristics, and risk of bias results

| Section | Detailed summary |
| --- | --- |
| **Study selection** | Total of 796 records identified through database searches. After removing duplicates and screening titles/abstracts, 100 full-text articles assessed. Of these, 45 rsfMRI studies were excluded for not measuring depressive symptoms and/or not examining associations with functional connectivity. An additional 7 studies were excluded for using non-resting-state modalities (task-based fMRI or anatomical MRI), 1 for conducting MRI of the breast, and 1 for including mixed cancer types. Also excluded: 15 conference abstracts/posters, 3 study protocols, and 1 review article. A final 27 studies met the eligibility criteria: 15 cross-sectional (56%) and 12 longitudinal (44%). Longitudinal studies included 7 cancer-treatment (6 pre/post chemotherapy; 1 pre-treatment to 12 months post-diagnosis) and 5 behavioural/cognitive interventions (mindfulness, physical exercise, mobile game, music stimulation, metacognitive training). |
| **Risk of bias** | None of the studies achieved the maximum score (9 stars) on the Newcastle–Ottawa Scale (NOS), with most (24/27, 89%) showing some risk of bias. Based on NOS-to-AHRQ conversion, 4 studies (15%) were rated good quality (low risk), 17 (63%) fair quality (moderate risk), and 6 (22%) poor quality (high risk). Across all studies, 44% (12/27) recruited BC participants from a single hospital or cancer centre, and 33% (9/27) did not report recruitment methods. Most studies (78%, 21/27) controlled for age, and 63% (17/27) adjusted for other relevant confounders (education, menopausal status, time since chemotherapy).<br>Regarding cross-sectional studies, 47% (7/15) did not report how breast cancer diagnosis/treatment information was obtained, and 1 study relied solely on self-report. Only 1 study (7%) provided a sample size justification. Most studies (75%, 13/15) reported >80% completion rates for depressive symptom and rsfMRI measures, and 94% (12/15) adequately described statistical analyses.<br>In longitudinal studies, 33% (4/12) used a control group from the same source population. For example, Kardan et al. [30] recruited a healthy control group of participants who had received negative mammograms from the same oncology centre as the BC participants. |

None of the 7 longitudinal studies examining pre-post chemotherapy changes verified breast diagnosis/treatment exposure, whereas all 5 longitudinal pre-post intervention studies confirmed program engagement (via attendance, interviews, monitoring tools). Over half (58%, 7/12) measured baseline depressive symptoms and FC, and 92% (11/12) had follow-up intervals ranging from 1 week to 12 months. However, 1 study [24] conducted same day pre/post scans to assess the immediate effects of a task-based fMRI music paradigm. Dropout/attrition rates were reported in 58% (7/12), with <20% loss to follow-up in each case.

**Participant characteristics**

Across the 27 rsfMRI studies reviewed, a total of 1,423 BC participants and 560 healthy controls were examined. Most studies (21/27, 78%) recruited middle-aged BC participants (mean ages > 45 years; range: 40.3 - 63.6), and the majority (23/27, 85%) included only non-metastatic disease (stage 0-III).

Cancer treatment exposure was heterogeneous (Supplementary Table S4). Common chemotherapy regimens included Doxorubicin–Cyclophosphamide–Paclitaxel; Cyclophosphamide–Methotrexate–Fluorouracil; and Docetaxel–Epirubicin–Cyclophosphamide. Some BC participants also received radiation therapy (12/27, 44%), hormonal therapy (11/27, 41%), and/or surgery (7/27, 26%). Most studies (20/27, 74%) did not report complete treatment details.

BC participants were typically described in studies based on treatment exposure or stage of cancer care. The most frequent group studied comprised newly diagnosed patients who had not yet received chemotherapy (BC-; 17 studies), of which six of these were longitudinal studies and reassessed participants post-chemotherapy. Nine studies included BC participants post-chemotherapy only, and another nine examined those who had received a combination of cancer treatment such as chemotherapy, radiation, and/or surgery (BC-C). Ten studies (38%) further divided BC participants by clinical symptoms or syndromes, including menopausal status [43], neuropathic pain [37], post-mastectomy pain, lymphedema, vestibulocerebellar ataxia [21], fatigue [27], cognitive impairment [38,41,42], or depression [21, 34-36].

Comparison group structures varied: 10 studies compared a single BC group to an healthy control group; one study compared a BC group to two healthy control groups; two studies compared BC participants with vs. without chemotherapy; five studies included all three participant groups (breast cancer participants with vs without chemotherapy vs healthy controls); and five employed single-cohort BC designs
