## Supplementary Table S5 Clinical Variables for "Investigating the link between depressive symptoms and resting-state brain connectivity in people with breast cancer: A systematic review"

**Supplementary Table S5.** Overview of the clinical variables of breast cancer participants from the included studies

| Study | Time since chemotherapy Mean (SD) months | Chemotherapy agents | Surgery | Radiation | Hormonal treatment |
|---|---|---|---|---|---|
| Bruno et al. 2012 | 64 | Y<br>Doxorubicin, cyclophosphamide, paclitaxel or docetaxel (79%)<br>Cyclophosphamide, methotrexate, fluorouracil (15%)<br>Doxorubicin, cyclophosphamide + cyclophosphamide, methotrexate, fluorouracil (6%) | Not reported | Y - 68% | Y - tamoxifen 56% |
| Bukkieva et al. 2022 | - | Y - chemotherapy agents and number of participants who received chemotherapy not reported | Y - Unilateral or bilateral Patey radical mastectomy (100%) | Y | Not reported |
| Chen et al. 2017 | NA - excluded participants who received chemotherapy | N | - | Y - 39% | Y – 20mg tamoxifen daily > 24m (100%) |
| Chen et al. 2020 | - | Y | - | - | - |

| | | | | | |
|---|---|---|---|---|---|
| Choghazardi et al. 2025 | - | - | Y – mastectomy | - | - |
| Donofry et al. 2022 | - | - | - | - | - |
| **Feng et al. 2020 [26]** | 0.38 since diagnosis - scheduled to commence chemotherapy | Y<br>Doxorubicin, cyclophosphamide, paclitaxel (62%)<br>Docetaxel, epirubicin, cyclophosphamide (38%) | Y – 69% | N | N |
| **Feng et al. 2020 [19]** | NA – scheduled to commence chemotherapy | Y<br>Post-operative:<br>Doxorubicin, cyclophosphamide, paclitaxel (65%)<br>Neoadjuvant:<br>Docetaxel, epirubicin, cyclophosphamide (29%)<br>Doxorubicin, cyclophosphamide, paclitaxel (1%) | Y | - | Y – 35% |
| Hampson et al. 2015 | 82 (35.4) since diagnosis | Y – 77% | Y – 100% | Y – 80% | Y – 55% |

| | | | | | |
|---|---|---|---|---|---|
| Harrison et al. 2021 | 68.4 (62.4) since primary treatment (surgery, chemotherapy, radiation) | Y<br>Doxorubicin, cyclophosphamide, paclitaxel or docetaxel (19%)<br>Cyclophosphamide, methotrexate, fluorouracil (5%)<br>Doxorubicin, cyclophosphamide (13%)<br>Cyclophosphamide, paclitaxel or docetaxel (11%)<br>Doxorubicin, cyclophosphamide, fluorouracil (1%) Epirubicin, cyclophosphamide, paclitaxel (1%) | - | Y – 69% | Y – 51% tamoxifen |
| Henneghan et al. 2022 | 39.2 (32.3) | Y – 100% | - | Y – 77% | Y – 69% |
| **Kardan et al. 2019** | NA – scheduled to commence chemotherapy | Y – 45% | Y | Y | - |
| Kesler et al. 2013 | NA – 58.8 (40.8) since chemotherapy/radiation | Y – 53%<br>Doxorubicin, cyclophosphamide, paclitaxel or docetaxel<br>Cyclophosphamide, methotrexate, fluorouracil<br>Doxorubicin, cyclophosphamide + cyclophosphamide, methotrexate fluorouracil | - | Y – 65% | Y – 55% |
| Kesler et al. 2017 | NA – included BC prior to any treatment (surgery, chemotherapy, radiation) - 1.2 (0.9) since diagnosis | N | N | N | - |

| | | | | | |
|---|---|---|---|---|---|
| Kesler et al. 2021 | 44.3 (43.5) since primary treatment | Y – 51% | - | Y – 40% | Y – 34% |
| Kim et al. 2018 | 34 (6) since diagnosis | Y | - | - | - |
| Liang et al. 2024 [35] | - | N | - | - | - |
| Liang et al. 2024 [36] | - | - | - | - | - |
| Liu et al. 2022 | - | - | - | - | - |
| Melis et al. 2023 | 25.6 (14.8) | Y – 100% | - | Y – 75% | Y – 73% endocrine therapy |
| **Phillips et al. 2022** | NA – scheduled to commence chemotherapy | Y<br>Anthracycline chemotherapy (65%) | Y – 100% | Y – 70% | Y<br>Hormone blockade (63%)<br>Selective estrogen receptor modulator (58%)<br>Other hormone suppressors (5%)<br>Aromatase inhibitors (51%)<br>Combination therapy (12%) |
| Shen et al. 2021 | - | Y<br>Anthracycline or taxane chemotherapy (50%) | - | - | - |
| Van der Gucht et al. 2020 | 18.96 since treatments | Y<br>Epirubicin, cyclophosphamide, paclitaxel (70%)<br>Fluorouracil, epirubicin, cyclophosphamide. Doxetaxel (15%)<br>Carboplatin + paclitaxel, epirubicin, cyclophosphamide (9%)<br>Docetaxel, cyclophosphamide (6%) | - | Y – 36% | Y – 79% tamoxifen citrate |
| Wolf et al. 2016 | Median 9.5 (range: 7- 34) | Y – 100% | - | - | - |

| | | | | | |
|---|---|---|---|---|---|
| **Yang et al. 2025** | NA – scheduled to commence chemotherapy | Y<br>Doxorubicin, cyclophosphamide, docetaxel (24%)<br>Epirubicin, cyclophosphamide, docetaxel (2%)<br>Paclitaxel, doxorubicin, cyclophosphamide (31%)<br>Doxorubicin, docetaxel (4%)<br>Docetaxel, carboplatin, trastuzumab, pyrotinib (16%)<br>Docetaxel, carboplatin, trastuzumab, pertuzumab (24%) | N | - | N – excluded patients receiving hormonal therapy |
| Zhou et al. 2022 | 40.3 (23.3) | Y<br>Docetaxel, cyclophosphamide (6%)<br>Fluorourracil, epirubicin, cyclophosphamide, docetaxel (11%)<br>Epirubicin, cyclophosphamide, doxetaxel (31%)<br>Neoadjuvant chemotherapy after surgery and doxetaxel, cyclophosphamide (2%) | Y -<br>Breast conserving surgery (19%)<br>Modified radical mastectomy (79%)<br>Simple mastectomy (11%) | N | N |

| | | | | | |
|---|---|---|---|---|---|
| Zhuang et al. 2024 | NA – scheduled to commence chemotherapy | Y<br>Doxorubicin, cyclophosphamide, paclitaxel (62%)<br>Docetaxel, epirubicin, cyclophosphamide (38%) | - | N | N |

*Note:* Studies in **bold** are longitudinal studies where chemotherapy was the primary exposure with follow-up over time. A dash (–) indicates unclear or unreported data, while NA means not applicable. Y = treatment received, N = treatment not received. Percentage values indicate the proportion of breast cancer participants who received each treatment type.
